# Synergistic Role of NK Cells and Monocytes in Promoting Atherogenesis in Severe COVID-19 Patients

**DOI:** 10.1101/2023.11.10.23298322

**Authors:** Manuja Gunasena, Mario Alles, Yasasvi Wijewantha, Will Mulhern, Emily Bowman, Janelle Gabriel, Aaren Kettelhut, Amrendra Kumar, Krishanthi Weragalaarachchi, Dhanuja Kasturiratna, Jeffrey C Horowitz, Scott Scrape, Sonal R Pannu, Shan-Lu Liu, Anna Vilgelm, Saranga Wijeratne, Joseph S Bednash, Thorsten Demberg, Nicholas T Funderburg, Namal P.M. Liyanage

**Affiliations:** Department of Microbial Infection and Immunity, College of Medicine, The Ohio State University; Department of Veterinary Bioscience, College of Veterinary Medicine, The Ohio State University; School of Health and Rehabilitation Sciences, College of Medicine, the Ohio State University; Department of pathology, College of Medicine, The Ohio State University; Department of Mathematics and Statistics, Northern Kentucky University, KY, Highland Heights, KY, USA; Department of Internal Medicine, College of Medicine, The Ohio State University; The Steve and Cindy Rasmussen Institute for Genomic Medicine, Abigail Wexner Research Institute at Nationwide Children’s Hospital, Columbus, Ohio; Department of Pediatrics, Baylor College of Medicine, Houston, TX; Dorothy M. Davis Heart and Lung Research Institute, College of Medicine, The Ohio State University.

## Abstract

Clinical data demonstrate an increased predisposition to cardiovascular disease (CVD) following severe COVID-19 infection. This may be driven by a dysregulated immune response associated with severe disease. Monocytes and vascular tissue resident macrophages play a critical role in atherosclerosis, the main pathology leading to ischemic CVD. Natural killer (NK) cells are a heterogenous group of cells that are critical during viral pathogenesis and are known to be dysregulated during severe COVID-19 infection. Their role in atherosclerotic cardiovascular disease has recently been described. However, the contribution of their altered phenotypes to atherogenesis following severe COVID-19 infection is unknown. We demonstrate for the first time that during and after severe COVID-19, circulating proinflammatory monocytes and activated NK cells act synergistically to increase uptake of oxidized low-density lipoprotein (Ox-LDL) into vascular tissue with subsequent foam cell generation leading to atherogenesis despite recovery from acute infection. Our data provide new insights, revealing the roles of monocytes/macrophages, and NK cells in COVID-19-related atherogenesis.

## Introduction

An increasing body of evidence suggests that coronavirus disease 2019 (COVID-19), caused by severe acute respiratory syndrome coronavirus 2 (SARS-CoV-2) predisposes affected individuals to CVDs including acute myocardial infarction and stroke^1^. Atherosclerosis is the main pathophysiological process underlying most cases of CVDs in humans^2^. It is a chronic inflammatory condition which can lead to coronary artery calcification and an overall prothrombotic state culminating in CVD pathology^3,4^. Monocytes and macrophages are central to the pathogenesis of atherosclerosis. Upon recruitment to the intima of arteries following lipid deposition, circulating monocytes differentiate into a broad spectrum of proinflammatory macrophages (M1 type), lipid-laden foam cells, and dendritic cells^5^.

Recent studies using coronary autopsy specimens from patients with COVID-19 revealed that infiltrating macrophages are infected by SARS-CoV-2, with foam cells, a hallmark of atherosclerosis at all stages of the disease, being more susceptible to SARS-CoV-2 infection than other macrophages. Furthermore, SARS-CoV-2 was found to induce a strong pro-atherogenic inflammatory response in both macrophages and foam cells, a phenomenon that was largely recapitulated in an *ex vivo* SARS-CoV-2 infection of human vascular explants, suggesting a potential mechanism for increased risk of atherosclerosis following COVID-19 infection^6^. Monocytes and macrophages are in constant direct and indirect (via soluble mediators) communication with several other immune cells.

During viral pathogenesis, one of the earliest cell types to arrive at the site of infection are NK cells which in partnership with monocytes/macrophages, trigger an immune response. A new understanding of the functions of monocytes and macrophages in health and diseases is emerging where NK cells play a crucial intermediary role^7^. The role of NK cells in “normal” atherosclerosis has been described previously. However, the phenotypic and functional associations of NK cells in COVID-19-associated atherosclerosis are not fully known.

For the first time, we demonstrate that in the peripheral blood of patients during and after severe COVID-19, monocytes and NK cells are activated and may influence the consumption of oxidized lipids from the circulation. This ongoing process results in the generation of foam cells in vascular tissue, suggestive of persistent atherogenesis even after recovery. *In vitro* experiments and correlational analyses have led to a mechanistic interpretation that suggests a direct crosstalk between activated NK cells and macrophages during COVID-19, leading to an increased uptake of Ox-LDL from the environment, subsequently resulting in foam cell generation and atherogenesis.

## Material and Methods

### Patient characteristics

Patients were enrolled in the Ohio State University Intensive Care Unit Registry (BuckICU, IRB: 2020H0175) at the Ohio State University Wexner Medical Center and the James Cancer Hospital and Solove Research Institute between April 2020 and January 2021. BuckICU is a multi-disciplinary biorepository project that collects human biospecimens and associated clinical data from critically ill patients admitted to the ICU (severe) with respiratory failure, defined as any increase in respiratory support (supplemental oxygen or need for invasive or non-invasive mechanical ventilation) above baseline. For this study, we analyzed samples from thirty-one critically ill patients with severe COVID-19 disease (SARS-CoV-2 RNA+) (n=31), between 28 and 69 years of age. Patients with current malignant diseases, ongoing immunomodulatory treatment before hospitalization, and known diabetes mellitus were excluded. Seventeen healthy controls (Control) that were SARS-CoV-2 immunoglobulin G (IgG) seronegative and symptom-free at sampling were recruited as controls, with ages ranging from 25 to 65 years old. To minimize inter-experimental variability and batch effects between patients and controls, all samples (n=77) were acquired, processed, and analyzed fresh during the period between April 2020 and January 2021. Another group of individuals at an age range of 24 to 60 were recruited after three months of recovery from COVID-19 infection (n=29) (Recovered) at the peak of the COVID-19 pandemic in the United States (August 2020). Supplementary Table 1 and Materials and Methods both provide more specific patient characteristics. To study the innate cell response, we stained fresh whole blood with a 20-color innate cell–focused flow cytometry panel. All clinical laboratory analyses, including serology and polymerase chain reaction (PCR) for SARS-CoV-2, were performed using clinical routine assays in the clinical laboratories at The Ohio State University Wexner Medical Center, Columbus, Ohio. We obtained approval by the Research Ethics Committee in The Ohio State University under following IRB protocols (2020H0175, 2020H0228, 2014H0467, 2020H0339) and all patients gave written informed consent. For detailed clinical information, see Supplementary Table 1.

### Flow cytometry staining and Data acquisition

Patients underwent a blood draw performed by a trained phlebotomist. Peripheral blood from healthy donors and patients was collected into sodium citrate tubes (BD, Franklin Lakes, NJ) and whole blood staining was performed. After collection, 120μL of whole blood was separated into polystyrene FACS (fluorescence-activated cell sorting) tubes and red blood cells were lysed using ACK buffer (Thermofisher, Waltham, Massachusetts) for 5 minutes followed by a wash with R10 media (RPMI-1640 supplemented with 10% FBS [fetal bovine serum], 2 mM L glutamine, 100 U/mL penicillin, and 100mg/mL streptomycin). After washing with phosphate-buffered saline (PBS), cells were pre-stained for viability exclusion using Zombie NIR Fixable Viability dye (BioLegend, San Diego, California) for 10 minutes, followed by a 20-minute incubation with a panel of directly conjugated monoclonal antibodies and TruStain diluted in equal parts of FACS buffer (PBS containing 0.1% sodium azide and 1% bovine serum albumin) and Brilliant Stain Buffer (BD Biosciences, San Jose, California). After staining, cells were fixed with 2% paraformaldehyde and were filtered by using filter caps for polystyrene FACS tubes before being acquired on a Cytek Aurora, a multispectral flow cytometry system with, 405-, 488-, and 640-nm lasers. Monoclonal antibodies used in the panel can be accessed in Supplementary Table 2.

### Flow cytometry data analysis

FCS files were exported from spectralflo software and imported into FlowJo v.10.6.2 for subsequent analysis with the use of the following plug-ins to visualize the data: DownSample (3.2), and t-distributed stochastic neighbor embedding (tSNE). First, the compensation matrix for the 20-color flow cytometry panel was generated to correct for fluorescence spillover emissions and applied to files. Dataset as such was used for the downstream analysis in both manual gating (Supplementary Fig.1) and automated analysis. For the automated analysis, events were first down-sampled from the CD45^+^ CD3^-^ CD19^-^ gate across all samples using Down Sample. Clinical parameter categorical values for each sample were added to downsampled populations as metadata to enable identification of these groups, and these were then concatenated for analysis. tSNE was run using all parameters from the panel except Zombie NIR (Live/Dead), BV711(CD19), FITC(CD45) and APC-Cy7 (CD3).

After generating the tSNE figures, manually gated immune cell subsets were overlaid to the figures to represent subsets from each cohort. (e.g., 17 “healthy controls,” 31“severe,” and 29 “recovered” patients). Computational analysis of flow cytometry data for monocytes were visualized by uniform manifold approximation and projection (UMAP) embedding. UMAP dimensionality reduction was done on the dataset using all parameters from the panel except Zombie NIR (Live/Dead), BV711(CD19) FITC(CD45) and APC-Cy7 (CD3). The data were projected in 2-dimensions using UMAP embeddings and clusters of major monocyte subsets were identified based on expression of lineage defining markers. We used R (version 4.2.1) with packages ggpubr, ggplot, psych, gridExtra, corrplot^8,9^ to generate figures.

### Measurements of inflammatory biomarkers, chemokines, and cytokines

The plasma collected from blood samples was stored at −20°C and batched until processing without a prior thaw. Using enzyme-linked immunosorbent assay (ELISA), the following inflammatory markers were measured (Supplementary Table 3) including: Tissue inhibitor of metalloproteinases-1 (TIMP-1), Tissue inhibitor of metalloproteinases-2 (TIMP-2), Monocyte chemotactic protein 1 (MCP-1), Lipoprotein-Associated Phospholipase A2 (LpPLA2), oxidized low-density lipoprotein (Ox-LDL). To detect plasma cytokine and chemokine levels, plasma samples were processed using the multiplexed ELISA-based platform Quantibody® Human Inflammation Array 3 (RayBiotech QAH-INF-3) in accordance with the manufacturer’s protocol. Raw optical data were analyzed using the manufacturer’s analysis tool to construct standard curves and determine absolute cytokine concentration.

### In-vitro Ox-LDL uptake experiment

An in-vitro experiment was conducted to investigate the role of activated NK cells in the uptake of Ox-LDL by macrophages. 5ml of venous blood samples were collected from severe COVID-19 patients (n=6) in sodium citrate tubes, and Peripheral blood mononuclear cells (PBMCs) were isolated using Ficoll gradient centrifugation. The isolation of NK cells was performed using the Miltenyi Biotec NK cell isolation kit (130-092-657). Subsequently, NK cells were activated *in vitro* through incubation with autologous plasma. Concurrently, PBMCs were allowed to rest and were incubated in Teflon-coated wells for up to six days. This incubation stimulated the development of monocyte-derived macrophages (MDMs) from peripheral blood monocytes^10^. After six days, these monocyte-derived macrophages were exposed to a medium containing Ox-LDL, which had been commercially manufactured and dye-tagged with Dil, to assess the absorption of Dil-Ox-LDL (10 ug/mL). This assessment was carried out after a four-hour incubation period in the presence or absence of plasma-activated NK cells. The cells were subsequently washed twice with PBS, and the intensity of DiI staining was measured using flow cytometry. To identify macrophages, markers such as CD45, CD3, CD19, and CD14 were utilized. For fluorescence microscopy analysis, the cells were seeded into 12-well plates and incubated at 37°C for an additional 24 hours. Images were acquired using an EVOS microscope, and the presence of Dil was detectable using an RFP filter (absorption peak: ∼554 nm, emission peak: ∼571 nm).

### scRNA-seq data analysis

scRNA-seq data were obtained from a published dataset^11^ The data file (in ‘rds’ format) downloaded from the cellxgene Data Portal (https://cellxgene.Cziscience.com/datasets) were read into R (version 4.2.1) session. Quality control was performed using Seurat (v3.1.5) to identify and filter out low-quality cells based on mitochondrial content and performing data preprocessing to prepare the data for downstream analysis. Cells with a percentage of mitochondrial genes greater than 10% were considered low quality and were filtered out. Prior to downstream analysis, the dataset was scaled and centered using “NormalizeData” and “ScaleData” functions. As the focus was on NK cells and monocytes and comparison between severe and control groups, specific subsets of scRNA-seq data set were separated and merged based on their metadata. The reanalysis included the standard Seurat workflow with “RunPCA“ and the number of Principle component analysis (PCA) dimensions used was 30, with a resolution parameter of 0.5. An elbow plot was generated to determine the optimal number of principal components for downstream analysis. UMAP plots were generated to visualize the cell clusters and identify potential subpopulations. After the cell clusters had been obtained, the Differentially expressed genes (DEG) in each cluster was computed with ‘FindAllMarkers’ (Supplementary Table 4)^12,13^. Marker genes were determined based on their average log-fold change (log_2_FC) and statistical significance (p-value and q-value).

Gene Ontology (GO) enrichment analysis was performed to gain insights into the biological processes associated with a set of genes of interest. The analysis was carried out using the clusterProfiler package in R (version 4.2.1)^13^. The significance threshold for enriched GO terms was set at a p-value cutoff of 0.05, and the Benjamini-Hochberg method was applied for multiple testing correction using GO terms with a q-value (adjusted p-value) below 0.05 were considered statistically significant. Cytoscape software (version 3.10.0) was used to visualize significant GO terms and integrate the GO terms to create a GO pathway network. For the DEGs, Kyoto Encyclopedia of Genes and Genomes (KEGG) pathway enrichment was performed for the top 200 DEGs. Enrichment analysis of the converted Entrez IDs was conducted to determine the significant KEGG pathways associated with the DEGs. The enrichKEGG function from the clusterProfiler package was utilized, specifying the gene IDs and the organism of interest (Homo sapiens, organism code ‘hsa’)^14^. This function identified overrepresented pathways based on a hypergeometric test. We employed the cnetplot function, to show the hub genes and pathways enriched.

### Statistical Analysis

Graphs and heatmaps were prepared using GraphPad Prism, version 9. Data were statistically analyzed used their bundled software. One-way ANOVA^15^ was used to compare three or more groups followed by Dunn’s multiple comparisons test for pairwise comparisons. Comparisons between two groups were performed using the Wilcoxon rank-sum test. To analyze the relationship between immune signatures and plasma biomarkers, Pearson correlation coefficient was calculated and plotted on a correlogram using R 4.2.1. Scatter plots showing linear relationships between data was created for significant correlations. Data visualization was achieved using functions from the Seurat, ggplot2, and pheatmap packages.

## Results

### Study design and cohort demographics

In this cross-sectional study, we assessed the immune profile in fresh whole blood samples from patients across three groups: 1) healthy donors: seventeen healthy age/sex-matched adults (n=17) who were SARS-CoV-2 immunoglobulin G (IgG) seronegative and asymptomatic at sampling as controls, 2) patients with severe COVID-19: thirty-one patients (n=31) with sampling on day one of intensive care unit admission for severe COVID-19 and respiratory failure, and 3) recovered patients: twenty-nine individuals with complete clinical recovery (n = 29) sampled three months after severe COVID-19 infection. Supplementary Table 1 and Materials and Methods both provide more specific patient characteristics. To minimize inter-experimental variability between patients and controls, all samples (n=77) were acquired, processed, and analyzed fresh between April 2020 and December 2020. Samples were analyzed using a 20-color spectral flow cytometer (Supplementary Table 1, gating strategy in Supplementary Fig.1).

### Temporal dynamics of innate immune signatures and dysregulations in severe COVID-19

We applied conventional flow cytometry and tSNE dimensionality reduction plots gated on CD45^+^CD3^-^CD19^-^ cells, which allowed for the identification and visualization of 10 innate immune cell populations (Fig.1A). The clustering algorithm allowed for an unbiased representation of these clusters during each stage of the disease. Among the dominant innate cell subsets that were focused on this study, we observed a significant increase in the frequencies of eosinophils (p<0.05) and neutrophils (p<0.05) among severe covid-19 patients (Fig.1B). While no differences were observed in the frequency of total monocytes between the control and severe groups, an overall increase in their frequencies was observed following recovery (p<0.05). Total NK cell frequencies were significantly decreased during severe infection and returned to basal levels after recovery, similar to previously published data^16^. We next analyzed the connectivity of cellular profiles and cytokine levels (detailed in Fig.2) in each study group. Heatmaps depicting correlation between 48 plasma cytokines/chemokines and 10 major innate immune cell subsets for the control, severe and recovered groups constructed based on Pearson correlation coefficient (Fig.1C). The correlation coefficients between those cell subsets were used to construct the density plot for each group. The density plot for Pearson correlation coefficients between three groups (control, severe, and recovered) comparing immune cell and cytokine data, provides a visual representation of the distribution of correlation coefficients for various pairs of variables between these groups (Fig.1D). Each line on the density plot represents one of the groups, and the plot can help reveal central tendency, spread and variability. A higher peak indicates a stronger central tendency toward certain correlation values. Visually comparing the three density lines, you can assess differences or similarities in the correlations between the groups. Comparing the three group’s density lines, we observe a greater overlap between severe and recovered group, compared to controls. Overall, the correlograms and density plots imply that the recovered donor group has a more similar distribution to severe patients than to the controls. PCA also revealed that three groups have differential cellular and cytokine profiles (Fig.1E). The volcano plot (Fig.1F) visualizes a sustained dysregulation of immune and biochemical parameters despite recovery from the disease. Overall, our data suggest that severe COVID-19 patients’ innate cell responses are dysregulated, with some immune perturbations persisting even after assumed viral clearance and recovery (up to 90 days post-infection).

### Differential inflammatory and cardiovascular biomarker profiles following severe COVID-19

Next, we analyzed 48 cytokines, chemokines, and plasma biomarkers related to cardiovascular risk, including those associated with the ‘cytokine storm’ and post-acute sequelae of SARS-CoV-2 infection, in plasma samples (Fig.2A). Interestingly, seven of 33 increased cytokine responses in the severe patients were also increased in the recovered population. Nine cytokines were decreased in severe patients and the recovered donors compared to levels in the healthy donors (Fig.2A). For a broader comparative analysis of proinflammatory protein profiles that exhibited differential expression among our study cohorts, we executed PCA on all the proteins identified in the plasma (Fig.2B). The control, severe, and recovered groups displayed distinct clustering, signifying significant dysregulation of inflammatory proteins both during and after severe COVID-19 infection. Interestingly, our findings reveal that metalloproteinase inhibitors, specifically TIMP-1 and TIMP-2, which are recognized for their pivotal roles in atherosclerosis, plaque development, and platelet aggregation^17^, were elevated in severe patients (Fig.2C and Fig.2D) and persisted even three months after recovery from severe COVID-19. We also found increased levels of MCP-1 (Fig.2E), which is known to mediate migration and infiltration of monocytes to tissues, in severe-COVID-19 patient’s plasma. We also found an increase in Interferon-gamma inducible Protein 10kDa (IP-10) (Supplementary Fig.2A), and decreased levels of CCL5/RANTES (Regulated on activation, normal T cell expressed and secreted) (Supplementary Fig.2B) and eotaxin (Supplementary Fig.2D) in severe patients compared to levels in the other groups. Levels of cardiometabolic and inflammatory biomarkers, including soluble CD14 (sCD14), lipopolysaccharide-binding protein (LBP), fatty acid binding protein 4 (FABP4), D-Dimer, C reactive protein (CRP), and tissue factor (Supplementary Fig.2), were elevated during severe disease, but levels in healthy and recovered participants were similar (Supplementary Fig.2).

Notably, CRP and D-dimer levels, indicative of COVID-19 severity, may correlate with complications like acute respiratory distress syndrome (ARDS) and cardiovascular events. Unexpectedly, levels of Ox-LDL and LpPLA2 were decreased in severe and recovered patients, compared to levels in healthy controls (Fig.2F-G). This surprising finding highlights a persistent disruption in mechanisms involving modified lipoprotein homeostasis during and after severe COVID-19 infection.

### Monocyte subset dynamics in COVID-19: Implications for immune dysregulation

Next, we examined monocyte subsets in our study populations. We observed elevated levels of CD14^++^CD16^−^ “classical” monocytes in the recovered donors’ whole blood compared to the other two groups (Fig.3A and B). In contrast, intermediate monocytes (CD14^++^CD16^+^) were increased in severe patients (Fig.3B), while the frequency of CD14^−^CD16^+^ “non-classical” monocytes was reduced in severe COVID-19 patients. Monocytes were visualized in a two-dimensional space using UMAP in Flowjo 10.9.0 (Fig.3C), which showed unique sub-clusters depending on the disease condition. Control and recovered groups had similar clustering patterns, distinct from the severe group, and some island clusters exhibited differences, suggesting varying features between these cohorts. No statistically significant changes were observed in CD27, CXCR3, and PD-1 receptor expression in classical and intermediate monocytes. However, the trend for decreased expression in PD-1 and increased CXCR3 expression in non-classical monocytes in the severe COVID-19 group suggests a more proinflammatory phenotype displayed by this subset (Fig.3D and Supplementary Fig.3C). We noted significant increases in CXCR5-expressing intermediate monocytes among our study populations compared to controls, with elevated frequencies persisting from severe disease into the recovery phase (Fig.3D and E). Similar monocyte/macrophage subsets have previously been reported in atherosclerotic lesions^18^. In contrast, severe COVID-19 patients exhibited low levels of intermediate and classical monocytes expressing tissue residency markers (e.g., CD69) in their peripheral circulation, and this reduction was also seen in the recovered group (p<0.05) (Fig.3D, and F). Our results indicate persistent dysregulation of circulating monocytes post-COVID-19 recovery, involving alterations in frequencies, subsets, and functional markers.

### Dysregulated NK cell phenotypes associated with COVID-19 infection

We and others have observed that COVID-19 infection alters total NK cell frequencies in peripheral blood^16,19^. Using a previously described gating strategy, CD56 and CD16 receptor expression were used to identify five distinct NK cell subsets^20^ (Fig. 4A and B). NK cell subsets clustered differently among the three groups. Overall, patients with severe illness had considerably lower frequencies of all five NK cell subsets (Fig.4D). Furthermore, these subsets showed noticeable differences between groups in terms of expression of activation and tissue homing markers (Supplementary Fig.4). In all NK subsets, CD69, a marker of activation and tissue residency^21^ was increased in individuals with severe COVID-19. Levels of CD69 on NK cell subsets was similar between healthy and recovered donors (Fig.4F). We also noted an increasing trend in HLA-DR expression during severe infection, suggesting NK cell activation during severe disease (Fig. 4G). Additionally, NKG2A expression (indicating an inhibitory NK phenotype)^22^ was decreased among NK subsets in severe COVID-19 patients (Supplementary Fig.4B), confirming NK cell activation during severe infection. We also noted significantly lower expression of CD27 in three of the NK subsets except CD56^dim^CD16^-^ and CD56^-^CD16^dim^ subsets in severe COVID-19 patients and recovered donors compared with controls (Fig.4E). CD27^+^ NK cells are particularly important in mediating NK cell /T cell interaction via the CD27-CD70 pathway^23^.

### Implications of activated NK cells and monocytes on Oxidized LDL levels and potential atherogenesis in severe COVID-19

In order to determine whether the changes we observed in innate cells were related to cardiometabolic biomarker levels, we performed Pearson correlational analysis to assess associations between plasma biomarker data acquired through ELISA and flow cytometry immune cell frequencies. As shown in Fig.2F and G, while most biomarkers returned to basal levels after recovery, Ox-LDL and LpPLA2 (which co-traffics Ox-LDL into tissue) levels remained decreased significantly even after recovery. Ox-LDL is recognized as a key player in the development of atherosclerosis (atherogenesis)^24^. We identified multiple correlations between immune cell data and plasma Ox-LDL (Fig.5A and B). We discovered a strong negative correlation between CD69^+^ NK cells and Ox-LDL levels (Fig.5C-D and Supplementary Fig.5A-D) as well as a negative correlation between Ox-LDL and activated (CD69^+^) total monocytes (Fig.5F), activated (CD69^+^) classical monocytes (Fig.5G) and intermediate monocytes (Fig.5H) among our severe COVID-19 patients. Monocytes migrate into the arterial wall and differentiate into macrophages, where these cells can internalize Ox-LDL and transform into foam cells^25^. Therefore, our findings suggest a novel mechanism underlying the reduction of Ox-LDL levels in severe COVID-19 patients, potentially driven by activated NK cells contributing to increased consumption of Ox-LDL from the circulation by MDMs, leading to increased foam cell formation and subsequent atherogenesis in COVID-19 patients.

### Transcriptional profiling and functional differentiation of monocytes and NK cells in severe COVID-19

We validated the phenotypic changes observed in flow cytometry and plasma biomarker assays using a publicly available single-cell RNA sequencing (scRNA-seq) dataset from Schulte-Schrepping et al.^11^ For both NK cells and monocytes, we identified 2307 and 1815 DEGs (adjusted p-value < 0.05 & positive log_2_FC) (Supplementary Fig.6A), selecting the top 200 DEGs for GO enrichment analysis (Supplementary Table 4 and 5). UMAP figures in Supplementary Fig.6B showcased immune cell annotations and clustering based on disease condition (control vs. severe COVID-19), with DEG analysis separately conducted for NK cells and monocytes. Heatmaps in Supplementary Fig.6C (monocytes) and Fig.6D (NK cells) provided an overview of transcriptional profile differences. Our analysis, incorporating the top 200 DEGs, revealed that transcriptional profiles phenotypically and functionally distinguished severe COVID-19 patients from uninfected counterparts. Volcano plots (Supplementary Fig.6D and E) displayed upregulated (red) and downregulated (green) genes. Our flowcytometry analysis revealed that severe patients and recovered donors both exhibited an increased presence of intermediate monocytes (as shown in Fig.2B). This difference was found to be statistically significant (p<0.05). Incidentally, our transcriptome analysis revealed an upregulation of *CD36*, a gene known to be more expressed in intermediate monocytes. *CD36* encodes a receptor for Ox-LDL, and it plays a role in binding and internalizing Ox-LDL. Additionally, our analysis predicted multiple epigenetic modifications occurring as a result of severe infection. This prediction was corroborated by a high number of DEGs associated with histone modification and DNA methylation. Examples of these genes include *H1-4, H1-3, H1-10, H1-2,* and *SETD2* (Supplementary Table 4). In monocytes, we observed significant upregulation in expression of *CXCL2*, *IL1b* and *NLRP3* other notable genes suggestive of a proinflammatory phenotype in monocytes. (Fig.6A & Supplementary Table 4). Similarly, in NK cells, we detected upregulation of genes indicative of migration, inflammation, and activation, including *ICAM3, TNFAIP3, CD69,* and *IL12RB1* (Fig.6B & Supplementary Table 4). Some genes like *ICAM3, TNFSF12, TNFSF1214, NLRP3* and *BRD2* were upregulated in both cell types (Supplementary Table 4).

### Pathway analysis reveals intriguing interplay between viral response and lipid storage regulation in macrophages, highlighting persistent changes in innate cells

Furthermore, we conducted GO enrichment analysis to explore the functional enrichment of the DEGs, and the KEGG database was used to group these differentially expressed genes into gene pathways. We utilized the GO database and considered GO terms with a p-value < 0.05 significantly enriched (Fig6C-D & Supplementary Table 5). Our analysis revealed significant enrichment in GO terms associated with lasting cellular epigenetic changes. This pattern was consistent in both NK cells and monocytes during severe infection. Both cell types showed enriched pathways related to dysregulated unfolded protein response, particularly regulation of endoplasmic reticulum (ER) stress. In monocytes, we found significant enrichment in GO terms like “leukocyte chemotaxis”, “myeloid leukocyte migration”, and “phagocytosis engulfment”, suggesting potential activation and differentiation of migratory, phagocytic macrophages. In NK cells, GO pathway analysis highlighted the importance of “defense response to virus” and “viral-induced Pattern recognition receptor (PRR) pathway signaling” in combating SARS-CoV-2 infection.

Interestingly, we observed an overlap between DEGs and enriched GO terms in NK cells and monocytes. Genes like *STAT*, *TNFAIP3*, and *NFKBIA* were differentially expressed and enriched in multiple GO terms. We used Cytoscape v3.10.0 to create network visualizations of enriched GO pathways for monocytes (Fig.6E) and NK cells (Fig.6F). The monocyte network has 57 nodes, while the NK cell network has 39 nodes, each representing a specific GO pathway. The edges between nodes indicate potential relationships between pathways based on shared genes. Lighter node colors indicate greater statistical significance for pathway enrichment. Connections between pathways in the network imply shared genes, suggesting possible crosstalk or cooperation in biological processes. Node color signifies pathway enrichment significance, and lighter shades indicate stronger significance. Edges represent shared genes, hinting at potential functional links, like crosstalk or cooperation between pathways due to shared genes in biological processes. In monocytes, a significant network emerges with interconnected pathways such as macrophage activation, phagocytosis, ER stress, acute inflammatory response, and lipid storage regulation. This network has the potential to fuel atherogenesis through immune system activation. A prominent connection arises in NK cells between defense response to viruses, PRR signaling, and stem cell differentiation regulation. This suggests that NK cells, when encountering viruses, adopt an activated migratory phenotype, potentially becoming dominant in circulation. Furthermore, the KEGG pathway analysis (Supplementary.Fig.6G-H) adds another layer of evidence to the notion of enduring alterations in innate cells and their potential impact on the post-acute consequences of SARS-CoV-2 infection. The amalgamation of differential gene expression and GO enrichment analysis offers a holistic understanding of the molecular terrain associated with severe COVID-19 and the enduring modifications in innate cells. This insight can potentially reveal valuable biomarkers and therapeutic targets for further exploration.

### Enhanced Ox-LDL uptake by monocyte-derived macrophages in the presence of autologous NK cells treated with plasma from severe COVID-19 patients

Based on our observations of innate immune responses and plasma biomarkers in vivo and analysis of published single-cell data, we have identified a significant correlation between multiple activated NK cell subsets and plasma Ox-LDL. Our findings strongly imply that these activated NK cells play a pivotal role in enhancing the uptake of circulating Ox-LDL. To further investigate the contribution of activated NK cells to atherogenesis and substantiate our hypothesis, we conducted an in vitro experiment to explore the impact of activated NK cells on Ox-LDL uptake by macrophages (Fig.7A).

We employed a method as outlined in one of our earlier publications. Frozen PBMCs from severe COVID-19 patients (n = 6) were used to isolate NK cells. These NK cells were subsequently incubated with autologous plasma. Concurrently, autologous MDMs were generated from the same patient’s PBMCs by allowing them to rest in Teflon-coated wells for up to six days. Following a six-day incubation period, we incubated these MDMs for four hours with commercially manufactured Dil-labeled Ox-LDL, both in the presence or absence of activated autologous NK cells. Interestingly, we observed significantly higher Dil-Ox-LDL uptake in the presence of activated NK cells (Wilcoxon signed rank-test; p<0.05 for all) (Fig.7B and C). The levels of Dil-Ox-LDL were quantified using flow cytometry, and this was further confirmed through EVOS microscope images (Fig.6D). Microscopic images were acquired to detect cells with internalized Dil-Ox-LDL using an RFP/TRITC filter set (absorption peak: ∼554 nm, emission peak: ∼571 nm). In summary, our findings collectively suggest that activated NK cells, in conjunction with other plasma factors, may be linked to increased Ox-LDL uptake and increased cardiovascular risk in individuals with severe COVID-19.

## Discussion

The COVID-19 pandemic has shed light on a range of health concerns beyond acute respiratory symptoms of the disease, including atherosclerotic cardiovascular disease. Given that evidence from genome-wide association studies, imaging techniques and clinical intervention studies confirm innate immune involvement in accelerating atherosclerosis^26^, it is likely that the immune perturbations following SARS-CoV-2 infection may be a key driver in COVID-19 associated CVDs. Our study provides mechanistic evidence to support the importance of unique macrophage-NK crosstalk during severe COVID-19 infection in promoting long-term atherogenesis. We show that in the proinflammatory environment of severe COVID-19, subsets of NK cells with altered phenotypes help macrophages take up modified lipids (Ox-LDL) and enhance foam cell generation. Our study also highlights a persistence of the pro-atherogenic potential following recovery from severe COVID-19, as evidenced by these innate immune alterations and disturbances in cardiometabolic biomarkers persisting up to three months after recovery.

Monocytes and macrophages play a crucial role in the development of atherosclerosis, both as mediators of chronic inflammation and plaque generation through lipid uptake^27^. Monocytes migrate from the bloodstream to the vessel wall, where they transform into tissue macrophages. These tissue macrophages subsequently convert into foam cells by taking up modified cholesterol, in particular Ox-LDL, initiating atherogenesis^28^. Our observations of significant increases in circulating intermediate monocytes in our severe COVID-19 patients (and remaining elevated post-recovery) suggests that atherogenesis may occur more readily during and after severe COVID-19 infection, as proinflammatory monocyte subsets such as these have a greater potential for trans-endothelial migration and vessel wall adhesion^5^. Increased CXCR5 expression in this monocyte subset among both severe COVID-19 patients and those who recovered further enhances their chemotactic potential towards inflamed vascular intimal tissue rich in its ligand CXCL13^29^. In addition, we noted elevated levels of other chemokine ligands like MCP-1^30^ in the plasma of severe COVID-19 patients. This may be a valid explanation to our observations of decreased CD69-expressing intermediate (tissue-resident) monocytes in the circulation of severe COVID-19 patients. Simultaneous reductions in Ox-LDL and LpPLA2 during severe infection, persisting even after recovery may indicate ongoing consumption of these atherogenic mediators. This may suggest that these intermediate monocytes leaving the circulation to transform into intimal macrophages may play a role in Ox-LDL consumption.

Contradicting evidence suggest that in COVID-19 patients, during early recovery, higher levels of Ox-LDL are likely to be observed^31^. However, these studies did not account for other risk factors of atherosclerosis and those which dysregulate lipid metabolism, including diabetes mellitus and hypertension. Considering the fact that we excluded majority of these factors suggest their exclusion may influence the outcome of our data which shows the onset of atherogenesis and role of Ox-LDL uptake in its early stages. Our analyses of transcriptome data from PBMCs of severe COVID-19 patients from external databases support our high-dimensional flowcytometry analyses, revealing enrichment in transcriptional pathways supporting chemotaxis, migration, phagocytosis, plasma membrane invagination, and engulfment, and upregulation of DEGs encoding macrophage scavenger receptors like CD36, SR-A and those involving lipid storage.

Monocytes/macrophages are the most investigated immune cells in atherosclerosis. However, there is now increasing preclinical and clinical evidence that NK cells play a significant role in its pathogenesis. Traditionally considered immune defenders against viruses^32,33^ and cancer^34^, this heterogenous group of immune cells are now increasingly recognized for their diverse roles, including those related to disease pathogenesis. In the case of human atherosclerosis, most of the current knowledge on NK cell involvement comes from observational studies on atherosclerotic lesions using immunohistochemistry. Bonaccorsi et al. characterized NK cells in atherosclerotic plaques of asymptomatic patients and found that these were enriched in CD56^bright^perforin^low^ NK cells, which also expressed tissue-resident markers such as CD69^35^.

While we and others have shown overall reductions in circulating NK cells during acute COVID-19^16^, we have also seen significantly increased expression of CD69 in these NK cells and their strong negative association with plasma Ox-LDL, suggesting that these NK cells may be linked to processes involving uptake of Ox-LDL from the circulation into tissue. Cytokines like TNFα, among the other proinflammatory cytokines that likely flood the circulation during the cytokine storm of acute severe COVID-19, are known to be key modulators of NK cell cytotoxicity, cytokine production, and adhesion molecule upregulation, and influences receptor expression, including activating receptors like NKG2D and CD69. As evidenced by our NK cell transcriptional analysis showing enrichment of the chemokine signaling pathway, these activated NK cells may be increasingly recruited to inflamed tissue to eliminate infected cells^36^. However, excessive NK cell activation can also promote tissue damage and inflammation.

Within this microenvironment, macrophages are reported to express NKG2D ligands (such as Major Histocompatibility Complex class I chain-related proteins A, MICA) suggesting the potential for direct macrophage-NK cell interactions. These interactions are known to promote macrophages to differentiate into proinflammatory M1 phenotype^37^.

Netea et al. demonstrated that, in addition to microbial stimuli, endogenous metabolites, such as Ox-LDL, that are abundant in the plaque microenvironment, train pro-atherogenic monocytes and macrophages, possibly via epigenetic modifications which may be the key driver leading to ongoing atherogenesis despite recovery from acute SARS-CoV-2 infection^38^. This would substantially increase future CVD risk in previously infected individuals. Analysis of single cell transcriptome data of monocytes and NK cells found enriched pathways related to histone modifications, crucial for epigenetic changes, further confirming the possibility of trained immunity following severe COVID-19 infection.

Within vascular intimal tissue, hydrolysis of Ox-LDL into oxidized fatty acids contribute to endothelial dysfunction, inflammation, and foam cell formation which may alter vascular architecture^39^. Progressive atherogenesis and vascular remodeling, reflected by persistent elevations in plasma TIMP-1 and TIMP-2 despite recovery^40^ is likely to contribute to the increased thromboembolic events documented following acute severe COVID-19 which has drawn significant scientific interest^41^. Indeed, Severe COVID-19 patients in our study showed a likely procoagulant state, evident from significantly elevated D-dimer levels, a marker of thromboembolic disease severity^42^. While the procoagulant state in COVID-19 is complex^43^, our study suggests that unique dysregulations in monocyte/macrophage and NK cell activity may contribute to thrombosis, by damaging the endothelium, promoting atherogenesis, and disrupting wall architecture. This interpretation aligns with the classical Virchow’s triad, which describes factors contributing to thrombosis. NK cell infiltration in particular may additionally damage the endothelium and disrupt wall architecture via perforin and granzyme B^44^ and elevate tissue factor levels (as seen in our severe COVID-19 patients), initiating the coagulation cascade by combining with activated factor VII^45^.

Our study is limited to the analysis of a small, cross-sectional cohort of individuals with severe COVID-19 infection. Furthermore, all samples were collected during the early phases of the COVID-19 pandemic, and thus our findings are pertinent only to the SARS-CoV-2 strains that circulated during that time in a pre-vaccination timeframe. Despite these limitations, our study provides a novel perspective on the pathophysiological events driven by monocytes and NK cells during severe COVID-19. These events ultimately contribute to atherogenesis and coagulopathy, leading to enduring cardiovascular and thromboembolic disease risks. The insights gained from this research pave the way for future therapeutic strategies aimed at mitigating COVID-related cardiovascular and thromboembolic events by targeting these mechanistic pathways.

## Acknowledgments

The authors gratefully acknowledge the support of all study individuals and their families. This work was partially supported by National Center for Advancing Translational Sciences (KL2TR002734 and UL1TR002733 to [J.S.B.]). We also, like to thank Sarah Karow and Gabrielle Swoope for their efforts in sample collection and clinical coordination.

## Author Contribution

M.G, Y.W and M.A performed experiments, data analysis and, wrote the manuscript. E.B, J.G and A.K, performed lipid biomarker experiments. A.V. and A.K, performed cytokine experiments and edited the manuscript. J.H, S.S, S.P, S.L and J.B coordinated clinical sample collection and edited the manuscript. D.K and K.W. supervised data analysis, S.W and M.G performed transcriptome data analysis, N.F supervised lipid biomarker experiments and wrote the manuscript, T.D, and N.L designed the study and wrote the manuscript.

## Data Availability

The data that support the findings of this study are available from the corresponding author upon reasonable request.

