## Supplemental Figures and Tables for "Synergistic Role of NK Cells and Monocytes in Promoting Atherogenesis in Severe COVID-19 Patients"

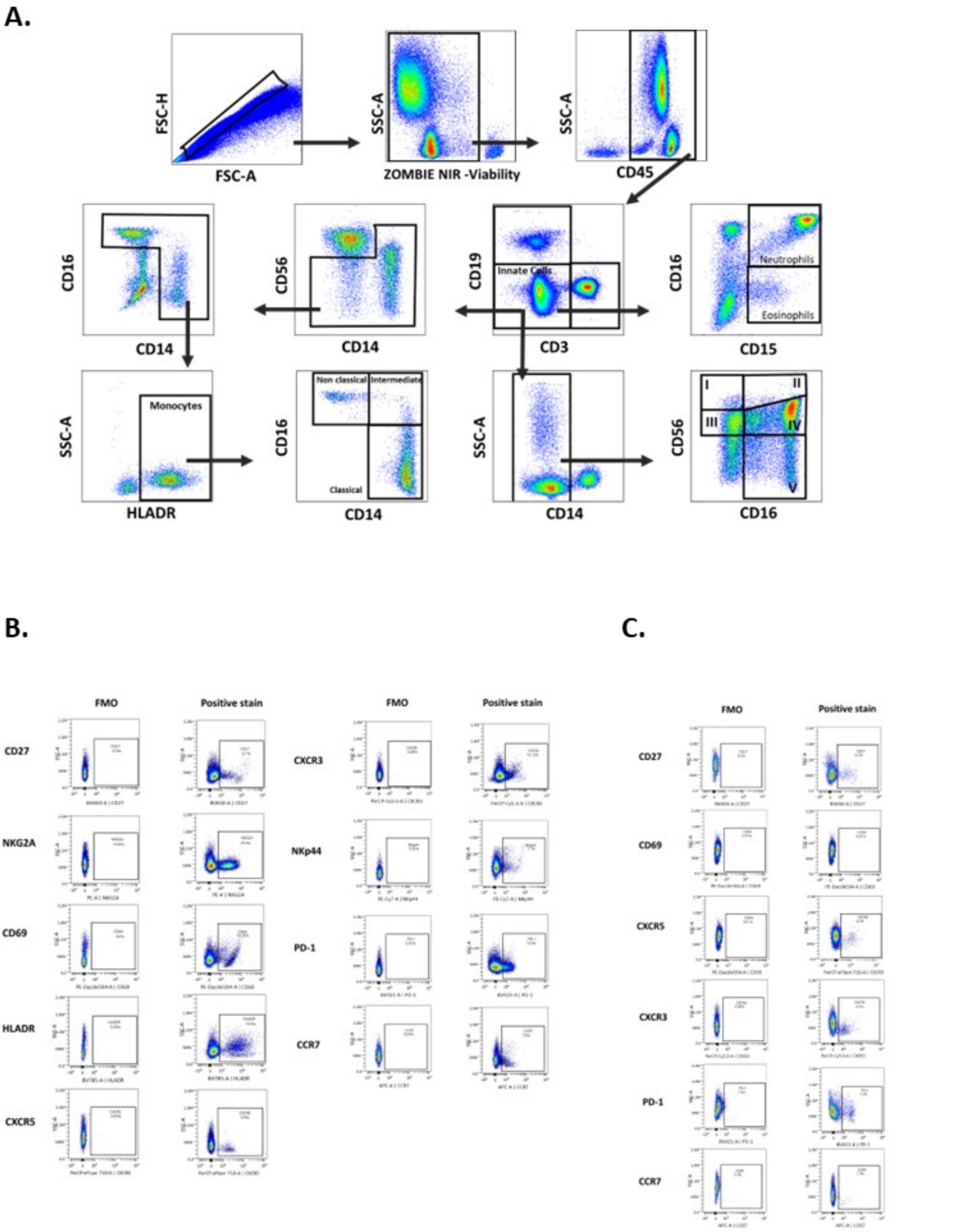

Supplementary Fig1: Representative pseudocolor plots for flow cytometry gating strategy (A) used to define cell populations whole blood staining. a) singlets. (b) Dead cells exclusion with ZOMBIE NIR viability dye. (c) Selection of CD45$^{+}$ cells. (d) Selection of CD3$^{-}$CD19$^{-}$ cells (e) gating of neutrophils CD16$^{+}$CD15$^{+}$ and eosinophils CD16$^{-}$CD15$^{+}$ (f) Exclusion of CD56$^{+}$ cells (g) Selection of CD14$^{+}$CD16$^{+}$ cells (h) Gating for total monocytes (i) Gating for three monocyte subsets based on CD14 and CD16 expression (j) Selection of CD14$^{-}$ cells (k) Gating for five NK cell subsets based on CD56 and CD16 expression. Representative pseudocolor plots for fluorescent minus one control for receptor expression in (B) NK cells and (C) monocytes

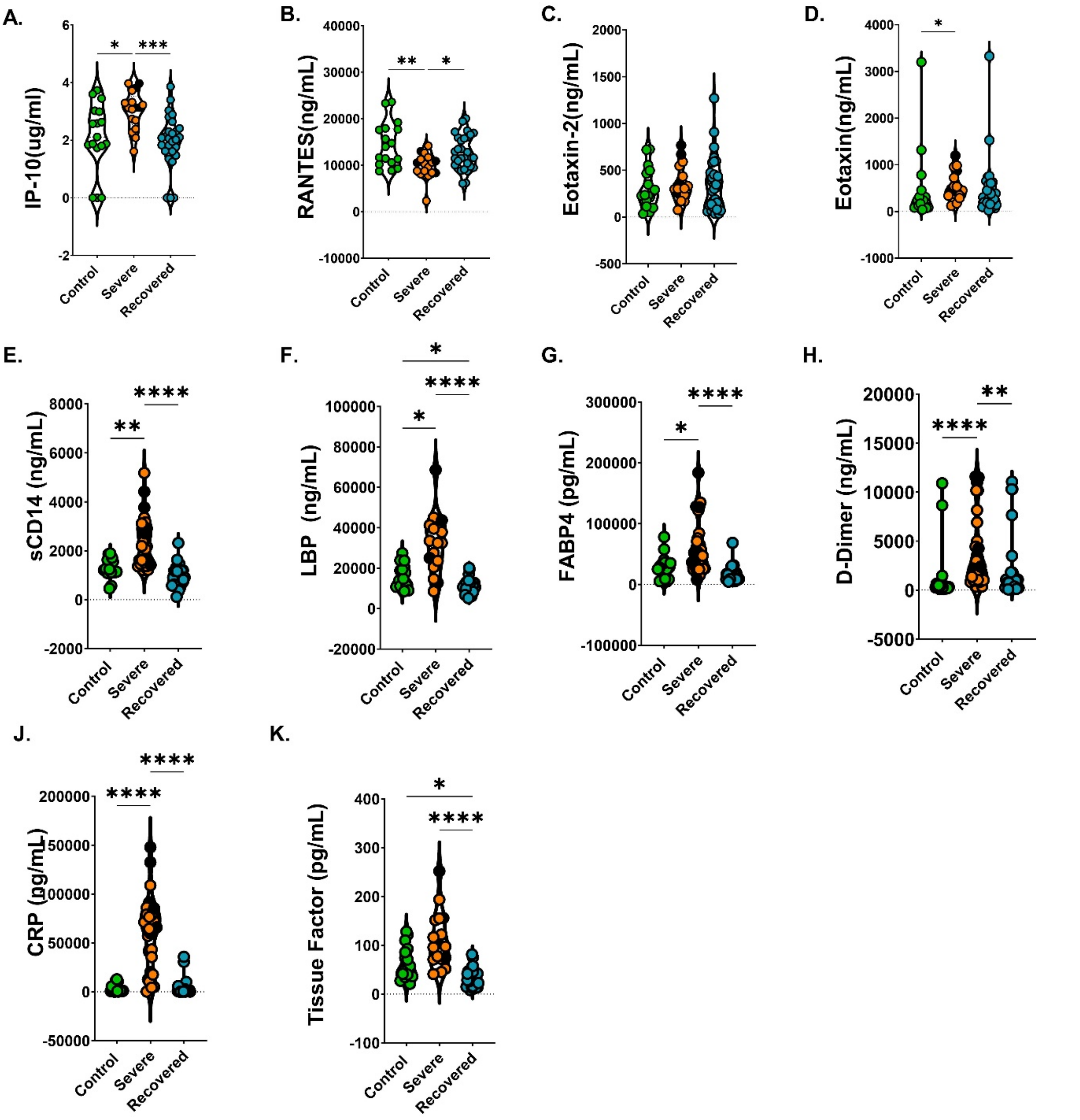

Supplementary Fig.2: Violin plots of plasma biomarker profiling of (A) IL-10 (B) RANTES (C) Eotaxin-2 (D) Eotaxin (E)sCD14 (F) LBP (G) FABP4 (H) D-Dimer (I) CRP (J) Tissue Factor determined in control, severe and recovered COVID-19 individuals.

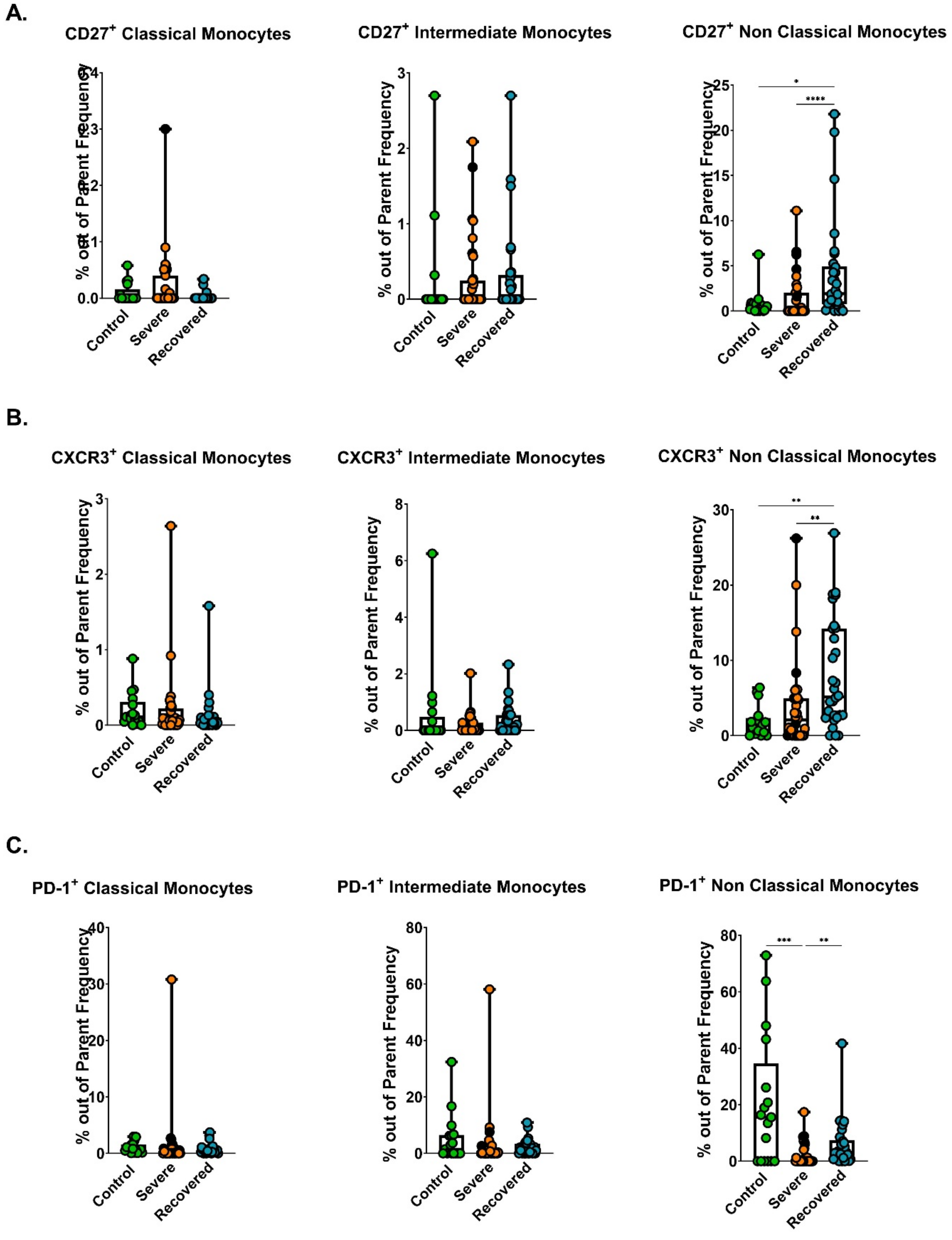

Supplementary Fig3: Monocyte cell subset frequency differences within viable CD45+ cells. (A) CD27 (B) CXCR3 (C) PD-1 expression differences in three monocyte subsets out of parent monocyte frequency between control, severe and recovered individuals. Differences between groups were calculated using Kruskal-Wallis test with Dunn's multiple comparison post-test. ***P < 0.001, **P < 0.01, and *P < 0.05.

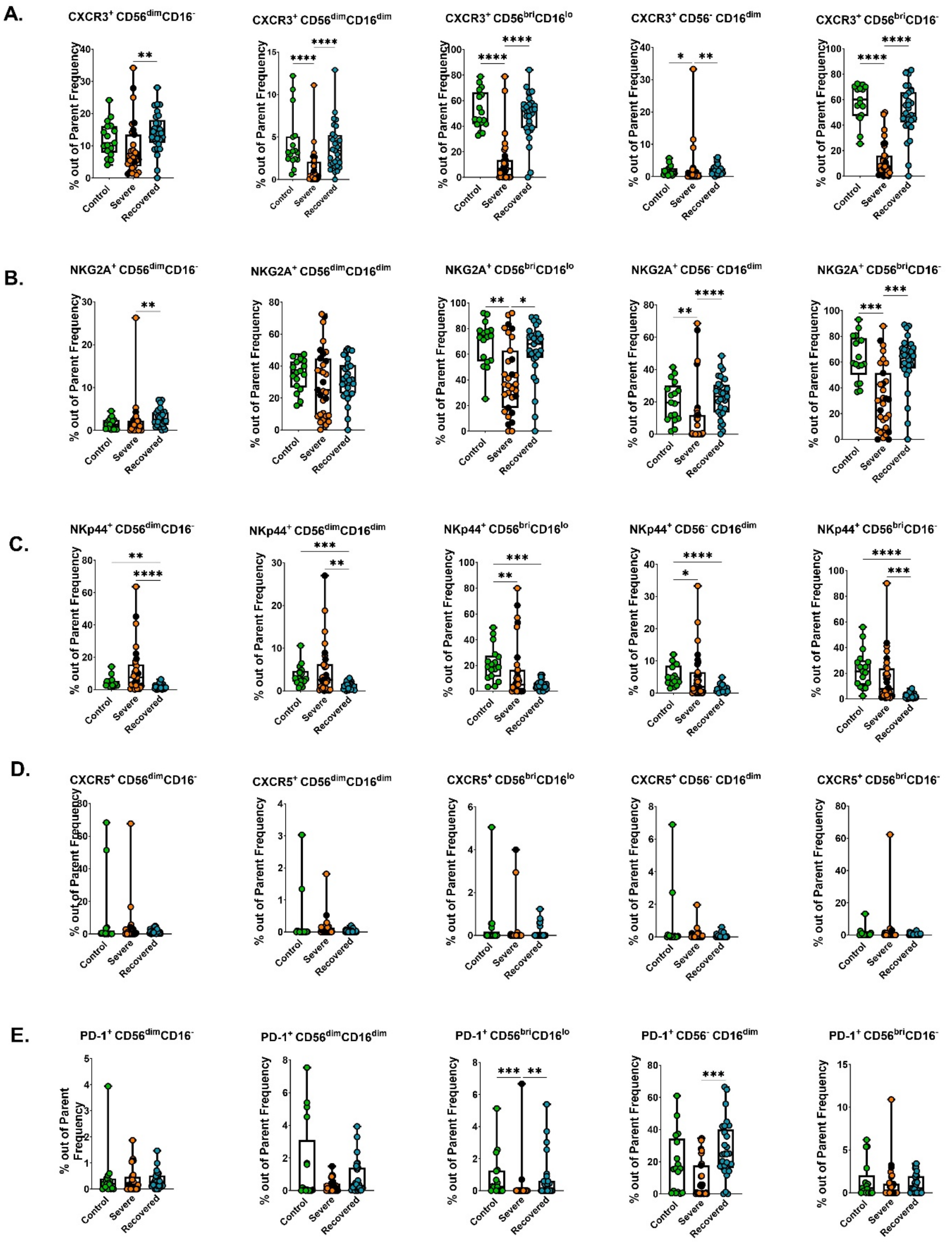

Supplementary Fig4: NK cell subset frequency differences within viable CD45+ cells. (A) CXCR3 (B) NKG2A (C) NKp44 (D)CXCR5 (E) PD-1 expression differences in five NK cell subsets out of parent NK frequency between control, severe and recovered individuals. Differences between groups were calculated using Kruskal-Wallis test with Dunn's multiple comparison post-test. ***P < 0.001, **P < 0.01, and *P < 0.05.

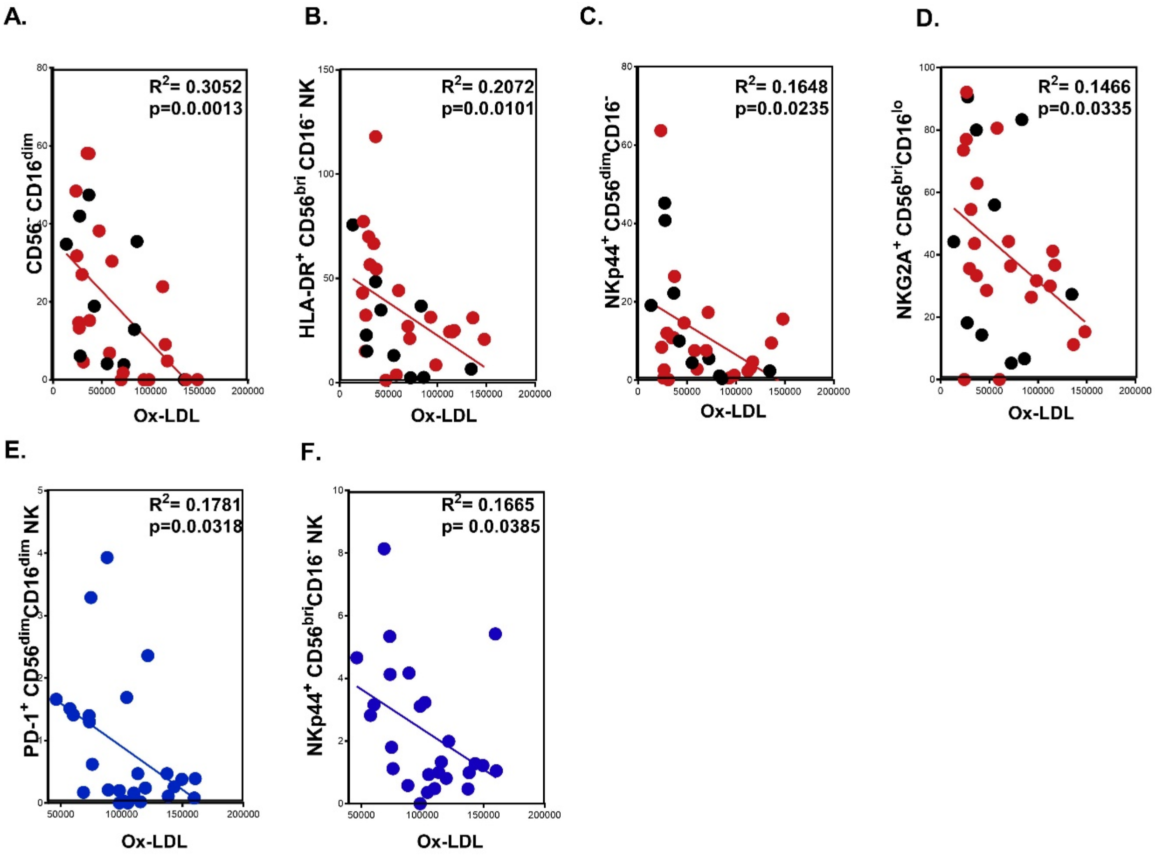

Supplementary Fig5: NK cell subset (A) $CD56^{-}CD16^{dim}$ (B) $HLADR^{+}CD56^{bri}CD16^{-}$ (C) $NKp44^{+}CD56^{dim}CD16^{-}$ (D) $NKG2A^{+}CD56^{bri}CD16^{lo}$ Correlations in severe COVID-19 with Ox-LDL. Black dots indicate deceased patients. (E) $PD\text{-}1^{+}CD56^{dim}CD16^{dim}$ (F) $NKp44^{+}$ $CD56^{bri}CD16^{-}$ Correlations in recovered individuals with Ox-LDL.

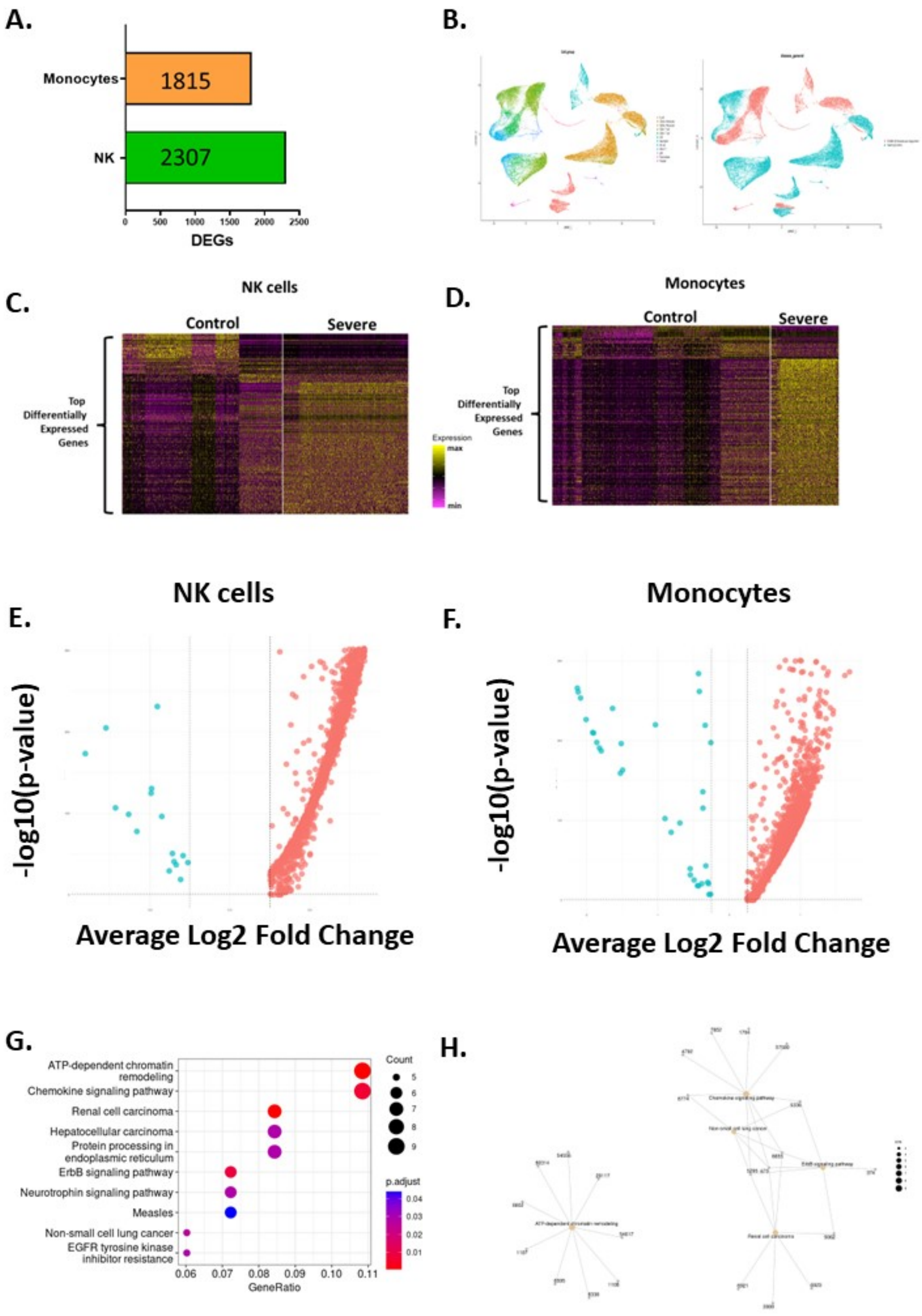

Supplementary Fig6: NK Cell subset (A) CD56$^{-}$CD16$^{dim}$ (B) HLADR$^{+}$CD56$^{bri}$CD16$^{-}$ (C) NKp44$^{+}$CD56$^{dim}$CD16$^{-}$ (D) NKG2A$^{+}$CD56$^{bri}$CD16$^{lo}$ Correlations in severe COVID-19 with Ox-LDL. Black dots indicate deceased patients. (E) PD-1$^{+}$CD56$^{dim}$CD16$^{dim}$ (F) NKp44$^{+}$ CD56$^{bri}$CD16$^{-}$ Correlations in recovered individuals with Ox-LDL.

**Supplementary Table 1. Patient demographic data**

| | Control | ICU | Recovered |
|---|---|---|---|
| **Sample number** | 17 | 31 | 29 |
| **Age range** | 25-65 | 28-69 | 24-60 |
| **Sex (M \|F)** | 9\|8 | 18\| 13 | 17\|12 |
| **BMI** | | 23.89 | |
| **Deaths** | - | 10 (32.25%) | - |
| **Race** | | | |
| White | 10 | 26 | 17 |
| Black | 5 | 5 | 8 |
| Asian | 2 | 0 | 4 |
| Other | 0 | 0 | 0 |
| **Vaccination Status** | 0 | 1 (3.22%) | 0 |
| **Bacterial infection** | - | 5(16.12%) | - |
| **Septic shock** | - | 19 (61.29%) | - |
| **Symptoms** | | | |
| Fever | - | 16 (51.61%) | - |
| Cough | - | 19 (61.29%) | - |
| Cough with sputum product | - | 5 (16.12%) | - |
| Sore throat | - | 0 | - |
| Runny nose | - | 0 | - |
| Wheezing | - | 2 (6.45%) | - |
| Chest Pain | - | 4 (12.9%) | - |
| Muscle aches | - | 5 (16.12%) | - |
| Joint pain | - | 0 | - |
| Fatigue/malaise | - | 13 (41.93%) | - |
| Shortness of breath | - | 30 (96.77%) | - |
| Swelling | - | 1 (3.22%) | - |
| Unable to walk | - | 2 (6.45%) | - |
| Headache | - | 3 (9.67%) | - |
| AMS/Confusion | - | 11 (35.48%) | - |
| Seizures | - | 1 (3.22%) | - |
| Syncope | - | 1 (3.22%) | - |
| Asmia | - | 1 (3.22%) | - |
| Abdominal pain | - | 1 (3.22%) | - |
| Nausea/Vomiting | - | 10 (32.25%) | - |
| Diarrhea | - | 10 (32.25%) | - |
| Conjunctivitis | - | 0 | - |
| Bleeding/hemorrhage | - | 0 | - |

**Supplementary Table 2. List of biomarkers assayed in plasma**

| | | | |
|---|---|---|---|
| BLC | IL-1ra | IL-12p40 | MIP-1β |
| Eotaxin | IL-2 | IL-12p70 | MIP-1d |
| Eotaxin-2 | IL-4 | IL-13 | PDGF-BB |
| G-CSF | IL-5 | IL-15 | RANTES |
| GM-CSF | IL-6 | IL-16 | TIMP-1 |
| I-309 | IL-6sR | IL-17 | TIMP-2 |
| ICAM-1 | IL-7 | MCP-1 | TNFα |
| IFNγ | IL-8 | MCSF | TNFβ |
| IL-1α | IL-10 | MIG | TNF RI |
| IL-1β | IL-11 | MIP-1α | TNF RII |

**Supplementary Table 3. List of antibodies for Flow Cytometric analyses.**

| Fluorochrome | Specificity | Clone | Company | Cat.No |
|---|---|---|---|---|
| FITC | CD45 | HI30 | Biolegend | 304006 |
| PE | NKG2a | Z199 | Beckman Coulter | IM3291U |
| PE/Dazzle 594 | CD69 | FN50 | Biolegend | 310942 |
| PE-Cy5 | CD4 | OKT4 | Biolegend | 317412 |
| PerCP-Cy5.5 | CXCR3 | G025H7 | Biolegend | 353714 |
| PerCP-eFluor 710 | CXCR5 | MU5UBEE | Thermo fisher | 46-9185-42 |
| PE-Cy7 | NKp44 | P44-8 | Biolegend | 325116 |
| APC | CCR7 | G043H7 | Biolegend | 353214 |
| APC-Cy7 | CD3 | SP34-2 | BD | 357757 |
| Brilliant Violet 421 | PD-1 | EH12.2H7 | Biolegend | 329920 |
| Super Bright 436 | CD56 | DREG56 | Thermo fisher | 62-0629-42 |
| BD Horizon BV480 | CD16 | 3G8 | BD | 1237529 |
| Pacific Orange | CD14 | TuK4 | Fisher Scientific | MCHD1430 |
| Brilliant Violet 570 | CD8 | RPA-T8 | Biolegend | 301038 |
| Brilliant Violet 605 | CD19 | HIB19 | Biolegend | 302243 |
| Brilliant Violet 650 | CD27 | O323 | Biolegend | 302827 |
| Brilliant Violet 711 | CD15 | HCD56 | Biolegend | 318336 |
| Brilliant Violet 750 | CD8 | 3A9 | BD | 747475 |
| Brilliant Violet 785 | HLADR | L243 | Biolegend | 307642 |

**Supplementary Table 4. Expression of the top two hundred DEGs in severe covid-19 compared with control.** In Excel file

#### Supplementary Table 5. GO pathways NK

##### Biological pathways

| ID | Description | GeneRatio | BgRatio | pvalue | p.adjust | qvalue | geneID | Count |
|---|---|---|---|---|---|---|---|---|
| GO:0006338 | chromatin remodeling | 18/181 | 411/21069 | 1.79E-08 | 4.59E-05 | 4.05E-05 | H1-4/H1-3/H1-2/ZBTB1/H1-10/BRD7/HP1BP3/CHD4/MIER1/KAT6B/CHD3/SPTY2D1/SMARCA2/DDX21/TSPYL2/SETD2/SMARCD1/INO80 | 18 |
| GO:0002753 | cytoplasmic pattern recognition receptor signaling pathway | 8/181 | 79/21069 | 3.94E-07 | 0.00033359 | 0.000294586 | TNFAIP3/RIOK3/NOP53/NFKBIA/SEC14L1/UFD1/ERBIN/CYLD | 8 |
| GO:0071824 | protein-DNA complex subunit organization | 14/181 | 306/21069 | 4.36E-07 | 0.00033359 | 0.000294586 | H1-4/H1-3/H1-2/H1-10/TAF10/HP1BP3/CENPC/KAT6B/SPTY2D1/BDP1/TSPYL2/MED23/SETD2/SMARCD1 | 14 |
| GO:0006354 | DNA-templated transcription elongation | 13/181 | 265/21069 | 5.20E-07 | 0.00033359 | 0.000294586 | ELOB/BCLAF1/ZNF326/PSIP1/ALYREF/KAT6B/CDK13/CCNK/KDM2A/CDK12/MED23/SETD2/SMARCD1 | 13 |
| GO:0034728 | nucleosome organization | 10/181 | 175/21069 | 2.87E-06 | 0.001298997 | 0.001147115 | H1-4/H1-3/H1-2/H1-10/HP1BP3/KAT6B/SPTY2D1/TSPYL2/SETD2/SMARCD1 | 10 |
| GO:0030098 | lymphocyte differentiation | 16/181 | 468/21069 | 3.04E-06 | 0.001298997 | 0.001147115 | ZBTB1/PIK3R1/RUNX3/BRD7/ITGA4/STAT3/NFIL3/DOCK2/PREX1/VSIR/SMARCA2/PLCG2/IRF2BP2/BRAF/SMARCD1/SOS2 | 16 |
| GO:0065004 | protein-DNA complex assembly | 12/181 | 269/21069 | 3.86E-06 | 0.00141547 | 0.00124997 | H1-4/H1-3/H1-2/H1-10/TAF10/HP1BP3/CENPC/KAT6B/SPTY2D1/BDP1/TSPYL2/MED23 | 12 |
| GO:0030968 | endoplasmic reticulum unfolded protein response | 7/181 | 82/21069 | 6.79E-06 | 0.002177004 | 0.001922463 | RACK1/PIK3R1/HSPA5/SELENOS/PPP1R15A/EIF2AK2/MBTPS1 | 7 |
| GO:0000018 | regulation of DNA recombination | 8/181 | 136/21069 | 2.35E-05 | 0.006343695 | 0.005601974 | H1-4/H1-3/H1-2/H1-10/EPC1/ALYREF/SETD2/ING3 | 8 |
| GO:0006334 | nucleosome assembly | 8/181 | 137/21069 | 2.47E-05 | 0.006343695 | 0.005601974 | H1-4/H1-3/H1-2/H1-10/HP1BP3/KAT6B/SPTY2D1/TSPYL2 | 8 |
| GO:0030217 | T cell differentiation | 12/181 | 336/21069 | 3.57E-05 | 0.008310195 | 0.007338546 | ZBTB1/PIK3R1/RUNX3/BRD7/STAT3/DOCK2/PREX1/VSIR/SMARCA2/BRAF/SMARCD1/SOS2 | 12 |
| GO:0018205 | peptidyl-lysine modification | 14/181 | 457/21069 | 4.31E-05 | 0.009200361 | 0.008124631 | H1-4/H1-3/H1-2/NSD3/BOD1L1/BRD7/TAF10/EPC1/BRD1/KAT6B/DDX21/SETD2/CBX4/ING3 | 14 |
| GO:0035966 | response to topologically incorrect protein | 9/181 | 198/21069 | 5.57E-05 | 0.01068313 | 0.009434031 | RACK1/PIK3R1/HSPA5/SELENOS/UFD1/CREBRF/PPP1R15A/EIF2AK2/MBTPS1 | 9 |
| GO:0030261 | chromosome condensation | 5/181 | 49/21069 | 6.22E-05 | 0.01068313 | 0.009434031 | H1-4/H1-3/H1-2/H1-10/ACIN1 | 5 |
| GO:0035967 | cellular response to topologically incorrect protein | 8/181 | 156/21069 | 6.25E-05 | 0.01068313 | 0.009434031 | RACK1/PIK3R1/HSPA5/SELENOS/UFD1/PPP1R15A/EIF2AK2/MBTPS1 | 8 |
| GO:0006352 | DNA-templated transcription initiation | 8/181 | 159/21069 | 7.15E-05 | 0.011452646 | 0.010113573 | ELOB/BCLAF1/TAF10/TFAM/GTF3C1/BDP1/ELOC/MED23 | 8 |
| GO:0006986 | response to unfolded protein | 8/181 | 163/21069 | 8.51E-05 | 0.011941507 | 0.010545276 | RACK1/PIK3R1/HSPA5/SELENOS/CREBRF/PPP1R15A/EIF2AK2/MBTPS1 | 8 |
| GO:1903706 | regulation of hemopoiesis | 14/181 | 491/21069 | 9.26E-05 | 0.011941507 | 0.010545276 | ZBTB1/PIK3R1/NFKBIA/RUNX3/BRD7/STAT3/BRD1/VSIR/KAT6B/SMARCA2/ACIN1/BRAF/SMARCD1/SOS2 | 14 |
| GO:0034620 | cellular response to unfolded protein | 7/181 | 123/21069 | 9.45E-05 | 0.011941507 | 0.010545276 | RACK1/PIK3R1/HSPA5/SELENOS/PPP1R15A/EIF2AK2/MBTPS1 | 7 |
| GO:0039532 | negative regulation of viral-induced cytoplasmic pattern recognition receptor signaling pathway | 3/181 | 11/21069 | 9.78E-05 | 0.011941507 | 0.010545276 | RIOK3/SEC14L1/UFD1 | 3 |
| GO:0098532 | histone H3-K27 trimethylation | 3/181 | 11/21069 | 9.78E-05 | 0.011941507 | 0.010545276 | H1-4/H1-3/H1-2 | 3 |
| GO:2000736 | regulation of stem cell differentiation | 6/181 | 88/21069 | 0.000111883 | 0.013039398 | 0.011514798 | STAT3/CHD4/EIF2AK2/CDK13/CHD3/CDK12 | 6 |
| GO:0018023 | peptidyl-lysine trimethylation | 5/181 | 58/21069 | 0.000140318 | 0.015642403 | 0.013813453 | H1-4/H1-3/H1-2/NSD3/SETD2 | 5 |
| GO:0039531 | regulation of viral-induced cytoplasmic pattern recognition receptor signaling pathway | 4/181 | 32/21069 | 0.000156981 | 0.016770835 | 0.014809946 | RIOK3/NOP53/SEC14L1/UFD1 | 4 |
| GO:0070423 | nucleotide-binding oligomerization domain containing signaling pathway | 4/181 | 34/21069 | 0.000199758 | 0.019211196 | 0.016964974 | TNFAIP3/NFKBIA/ERBIN/CYLD | 4 |
| GO:1900101 | regulation of endoplasmic reticulum unfolded protein response | 4/181 | 34/21069 | 0.000199758 | 0.019211196 | 0.016964974 | RACK1/PIK3R1/HSPA5/PPP1R15A | 4 |
| GO:0002244 | hematopoietic progenitor cell differentiation | 7/181 | 139/21069 | 0.000202302 | 0.019211196 | 0.016964974 | ZBTB1/SSBP3/SBDS/EIF2AK2/BRAF/ZBTB24/SOS2 | 7 |
| GO:0002221 | pattern recognition receptor signaling pathway | 9/181 | 236/21069 | 0.000211164 | 0.019336566 | 0.017075685 | TNFAIP3/RIOK3/NOP53/NFKBIA/SEC14L1/UFD1/ERBIN/CYLD/PLCG2 | 9 |
| GO:0035872 | nucleotide-binding domain, leucine rich repeat containing receptor signaling pathway | 4/181 | 35/21069 | 0.000224029 | 0.019807239 | 0.017491325 | TNFAIP3/NFKBIA/ERBIN/CYLD | 4 |
| GO:0034976 | response to endoplasmic reticulum stress | 10/181 | 295/21069 | 0.00024503 | 0.020941885 | 0.018493306 | RACK1/PIK3R1/SELENOK/HSPA5/SELENOS/UFD1/PPP1R15A/EIF2AK2/MAN1A1/MBTPS1 | 10 |
| GO:0006310 | DNA recombination | 11/181 | 354/21069 | 0.000255405 | 0.021124436 | 0.018654512 | H1-4/H1-3/H1-2/H1-10/EPC1/RAD21/ALYREF/SETD2/ING3/SUPV3L1/INO80 | 11 |
| GO:0043484 | regulation of RNA splicing | 8/181 | 193/21069 | 0.000271741 | 0.021773223 | 0.019227441 | PIK3R1/CCNL1/TAF10/RBM38/ZNF326/QKI/YTHDC1/CDK12 | 8 |
| GO:0051607 | defense response to virus | 11/181 | 361/21069 | 0.000301777 | 0.02279756 | 0.02013201 | TNFAIP3/RIOK3/SELENOK/NOP53/DDIT4/SEC14L1/UFD1/EIF2AK2/IFI6/DDX21/SETD2 | 11 |
| GO:0140546 | defense response to symbiont | 11/181 | 362/21069 | 0.000308948 | 0.02279756 | 0.02013201 | TNFAIP3/RIOK3/SELENOK/NOP53/DDIT4/SEC14L1/UFD1/EIF2AK2/IFI6/DDX21/SETD2 | 11 |
| GO:0030522 | intracellular receptor signaling pathway | 10/181 | 304/21069 | 0.000311199 | 0.02279756 | 0.02013201 | TNFAIP3/RIOK3/NOP53/NFKBIA/SEC14L1/UFD1/ERBIN/STAT3/CNOT1/CYLD | 10 |
| GO:0032786 | positive regulation of DNA-templated transcription, elongation | 5/181 | 70/21069 | 0.000341105 | 0.024294253 | 0.021453707 | ALYREF/CDK13/CCNK/CDK12/MED23 | 5 |
| GO:0006302 | double-strand break repair | 10/181 | 315/21069 | 0.000411755 | 0.028533484 | 0.025197276 | PAXX/BRD7/EPC1/RAD21/SMARCA2/KDM2A/SETD2/ING3/SMARCD1/INO80 | 10 |
| GO:0045580 | regulation of T cell differentiation | 8/181 | 214/21069 | 0.000541166 | 0.034201923 | 0.030202946 | ZBTB1/RUNX3/BRD7/VSIR/SMARCA2/BRAF/SMARCD1/SOS2 | 8 |
| GO:1900102 | negative regulation of endoplasmic reticulum unfolded protein response | 3/181 | 19/21069 | 0.000546024 | 0.034201923 | 0.030202946 | RACK1/HSPA5/PPP1R15A | 3 |
| GO:0039528 | cytoplasmic pattern recognition receptor signaling pathway in response to virus | 4/181 | 44/21069 | 0.000546911 | 0.034201923 | 0.030202946 | RIOK3/NOP53/SEC14L1/UFD1 | 4 |
| GO:0045910 | negative regulation of DNA recombination | 4/181 | 44/21069 | 0.000546911 | 0.034201923 | 0.030202946 | H1-4/H1-3/H1-2/H1-10 | 4 |
| GO:0045663 | positive regulation of myoblast differentiation | 4/181 | 45/21069 | 0.000596273 | 0.034823282 | 0.030751654 | RIPOR2/BRD7/SMARCA2/SMARCD1 | 4 |
| GO:0032434 | regulation of proteasomal ubiquitin-dependent protein catabolic process | 7/181 | 167/21069 | 0.000613945 | 0.034823282 | 0.030751654 | ELOB/RACK1/NOP53/RNF19A/BAG5/ZFAND2A/N4BP1 | 7 |
| GO:0097193 | intrinsic apoptotic signaling pathway | 10/181 | 332/21069 | 0.000619448 | 0.034823282 | 0.030751654 | RACK1/PIK3R1/SELENOK/BCLAF1/DDIT4/SELENOS/PPP1R15A/CYLD/IFI6/BAG5 | 10 |
| GO:0006368 | transcription elongation by RNA polymerase II promoter | 6/181 | 121/21069 | 0.000628495 | 0.034823282 | 0.030751654 | ELOB/CDK13/CCNK/CDK12/MED23/SETD2 | 6 |
| GO:0032784 | regulation of DNA-templated transcription elongation | 6/181 | 121/21069 | 0.000628495 | 0.034823282 | 0.030751654 | ZNF326/ALYREF/CDK13/CCNK/CDK12/MED23 | 6 |
| GO:0032495 | response to muramyl dipeptide | 3/181 | 20/21069 | 0.000638336 | 0.034823282 | 0.030751654 | TNFAIP3/NFKBIA/ERBIN | 3 |
| GO:0070816 | phosphorylation of RNA polymerase II C-terminal domain | 3/181 | 21/21069 | 0.000740038 | 0.039530346 | 0.034908357 | CDK13/CCNK/CDK12 | 3 |
| GO:0070972 | protein localization to endoplasmic reticulum | 5/181 | 84/21069 | 0.000789948 | 0.041335235 | 0.036502213 | HSPA5/SPCS3/PPP1R15A/ZFAND2A/MAN1A1 | 5 |
| GO:0006282 | regulation of DNA repair | 8/181 | 230/21069 | 0.000866291 | 0.04271438 | 0.037720105 | BRD7/TAF10/EPC1/SMARCA2/SETD2/ING3/SMARCD1/INO80 | 8 |
| GO:0006984 | ER-nucleus signaling pathway | 4/181 | 50/21069 | 0.000891417 | 0.04271438 | 0.037720105 | HSPA5/SELENOS/PPP1R15A/MBTPS1 | 4 |
| GO:0032479 | regulation of type I interferon production | 6/181 | 130/21069 | 0.000915298 | 0.04271438 | 0.037720105 | RIOK3/UFD1/REL/CYLD/SETD2/PLCG2 | 6 |
| GO:0032606 | type I interferon production | 6/181 | 130/21069 | 0.000915298 | 0.04271438 | 0.037720105 | RIOK3/UFD1/REL/CYLD/SETD2/PLCG2 | 6 |
| GO:0009615 | response to virus | 12/181 | 479/21069 | 0.000925523 | 0.04271438 | 0.037720105 | TNFAIP3/RIOK3/CXCR4/SELENOK/NOP53/DDIT4/SEC14L1/UFD1/EIF2AK2/IFI6/DDX21/SETD2 | 12 |
| GO:2001022 | positive regulation of response to DNA damage stimulus | 7/181 | 180/21069 | 0.000954356 | 0.04271438 | 0.037720105 | BCLAF1/BRD7/EPC1/SMARCA2/ING3/SMARCD1/INO80 | 7 |
| GO:0045047 | protein targeting to ER | 4/181 | 51/21069 | 0.000960856 | 0.04271438 | 0.037720105 | HSPA5/SPCS3/ZFAND2A/MAN1A1 | 4 |
| GO:0033962 | P-body assembly | 3/181 | 23/21069 | 0.000973059 | 0.04271438 | 0.037720105 | DYNC1H1/CNOT1/NBDY | 3 |
| GO:0080182 | histone H3-K4 trimethylation | 3/181 | 23/21069 | 0.000973059 | 0.04271438 | 0.037720105 | H1-4/H1-3/H1-2 | 3 |
| GO:0016072 | rRNA metabolic process | 9/181 | 292/21069 | 0.000982897 | 0.04271438 | 0.037720105 | RIOK3/TRIR/NOP53/SBDS/KRR1/BUD23/GTF3C1/DDX21/XRN1 | 9 |
| GO:0042113 | B cell activation | 10/181 | 354/21069 | 0.001010127 | 0.042761468 | 0.037761687 | IGHA1/IGLC3/TNFAIP3/ZBTB1/IGKC/PIK3R1/IGLC2/ITGA4/PLCG2/IRF2BP2 | 10 |
| GO:0050688 | regulation of defense response to virus | 5/181 | 89/21069 | 0.00102613 | 0.042761468 | 0.037761687 | TNFAIP3/RIOK3/SELENOK/SEC14L1/UFD1 | 5 |
| GO:0032968 | positive regulation of transcription elongation by RNA polymerase II | 4/181 | 52/21069 | 0.001034014 | 0.042761468 | 0.037761687 | CDK13/CCNK/CDK12/MED23 | 4 |
| GO:0070431 | nucleotide-binding oligomerization domain containing 2 signaling pathway | 3/181 | 24/21069 | 0.001105075 | 0.044974811 | 0.03971624 | TNFAIP3/NFKBIA/ERBIN | 3 |
| GO:2000781 | positive regulation of double-strand break repair | 5/181 | 92/21069 | 0.001190882 | 0.047016293 | 0.041519027 | BRD7/EPC1/SMARCA2/ING3/SMARCD1 | 5 |
| GO:1903573 | negative regulation of response to endoplasmic reticulum stress | 4/181 | 54/21069 | 0.001191911 | 0.047016293 | 0.041519027 | RACK1/HSPA5/SELENOS/PPP1R15A | 4 |
| GO:0016573 | histone acetylation | 7/181 | 189/21069 | 0.001266486 | 0.048863837 | 0.043150552 | BRD7/TAF10/EPC1/BRD1/KAT6B/DDX21/ING3 | 7 |
| GO:0072599 | establishment of protein localization to endoplasmic reticulum | 4/181 | 55/21069 | 0.001276863 | 0.048863837 | 0.043150552 | HSPA5/SPCS3/ZFAND2A/MAN1A1 | 4 |

#### Molecular function

| ID | Description | GeneRatio | BgRatio | pvalue | p.adjust | qvalue | geneID | Count |
|---|---|---|---|---|---|---|---|---|
| GO:0031492 | nucleosomal DNA binding | 5/185 | 41/20758 | 3.08E-05 | 0.00565659 | 0.00510159 | H1-4/H1-3/H1-2/H1-10/CHD4 | 5 |
| GO:0042393 | histone binding | 11/185 | 277/20758 | 4.06E-05 | 0.00565659 | 0.00510159 | BRD7/BRD1/CHD4/KAT6B/CHD3/SPTY2D1/SMARCA2/TSPYL2/CBX4/ING3/INO80 | 11 |
| GO:0016887 | ATP hydrolysis activity | 13/185 | 388/20758 | 4.76E-05 | 0.00565659 | 0.00510159 | HSPA5/DDX24/CHD4/KIF2A/VPS4B/CHD3/SMARCA2/DDX21/RFC1/ACIN1/SUPV3L1/ASCC3/INO80 | 13 |
| GO:0031491 | nucleosome binding | 6/185 | 75/20758 | 5.59E-05 | 0.00565659 | 0.00510159 | H1-4/H1-3/H1-2/H1-10/HP1BP3/CHD4 | 6 |
| GO:0001221 | transcription coregulator binding | 7/185 | 117/20758 | 8.66E-05 | 0.007016416 | 0.006327996 | ELOB/JUND/RUNX3/CHD4/TFAM/ZNF644/ELOC | 7 |
| GO:0008094 | ATP-dependent activity, acting on DNA | 7/185 | 127/20758 | 0.000144855 | 0.009777683 | 0.008818339 | CHD4/CHD3/SMARCA2/RFC1/SUPV3L1/ASCC3/INO80 | 7 |
| GO:0008353 | RNA polymerase II CTD heptapeptide repeat kinase activity | 3/185 | 14/20758 | 0.000235825 | 0.013644164 | 0.012305458 | CDK13/CCNK/CDK12 | 3 |
| GO:0031490 | chromatin DNA binding | 6/185 | 122/20758 | 0.000794457 | 0.040219408 | 0.036273254 | H1-4/H1-3/H1-2/H1-10/STAT3/CHD4 | 6 |
| GO:0004386 | helicase activity | 7/185 | 177/20758 | 0.001070503 | 0.047953746 | 0.043248732 | DDX24/CHD4/CHD3/SMARCA2/DDX21/SUPV3L1/ASCC3 | 7 |
| GO:0001222 | transcription corepressor binding | 4/185 | 52/20758 | 0.001184043 | 0.047953746 | 0.043248732 | ELOB/RUNX3/ZNF644/ELOC | 4 |

#### Cellular components

| ID | Description | GeneRatio | BgRatio | pvalue | p.adjust | qvalue | geneID | Count |
|---|---|---|---|---|---|---|---|---|
| GO:1904949 | ATPase complex | 9/187 | 125/22283 | 1.13E-06 | 0.00033714 | 0.000293165 | BRD7/CHD4/VPS4B/CHD3/SMARCA2/DDX21/ING3/SMARCD1/INO80 | 9 |
| GO:0000786 | nucleosome | 9/187 | 137/22283 | 2.42E-06 | 0.000350105 | 0.000304439 | H1-4/H1-3/H1-2/H1-10/H2AC20/HP1BP3/EPC1/KAT6B/ING3 | 9 |
| GO:0044815 | DNA packaging complex | 10/187 | 183/22283 | 3.51E-06 | 0.000350105 | 0.000304439 | H1-4/H1-3/H1-2/H1-10/H2AC20/HP1BP3/EPC1/RAD21/KAT6B/ING3 | 10 |
| GO:0032993 | protein-DNA complex | 11/187 | 234/22283 | 4.86E-06 | 0.000363604 | 0.000316177 | H1-4/H1-3/H1-2/H1-10/JUND/H2AC20/TAF10/HP1BP3/EPC1/KAT6B/ING3 | 11 |
| GO:0070603 | SWI/SNF superfamily-type complex | 8/187 | 121/22283 | 8.49E-06 | 0.000507766 | 0.000441536 | BRD7/CHD4/CHD3/SMARCA2/DDX21/ING3/SMARCD1/INO80 | 8 |
| GO:0071735 | IgG immunoglobulin complex | 3/187 | 10/22283 | 6.68E-05 | 0.002532944 | 0.00220256 | IGHA1/IGLC3/IGKC | 3 |
| GO:0071745 | IgA immunoglobulin complex | 3/187 | 10/22283 | 6.68E-05 | 0.002532944 | 0.00220256 | IGHA1/IGLC3/IGKC | 3 |
| GO:0008023 | transcription elongation factor complex | 5/187 | 51/22283 | 6.78E-05 | 0.002532944 | 0.00220256 | ELOB/CDK13/CCNK/ELOC/CDK12 | 5 |
| GO:0019908 | nuclear cyclin-dependent protein kinase holoenzyme complex | 3/187 | 11/22283 | 9.13E-05 | 0.003034021 | 0.002638279 | CDK13/CCNK/CDK12 | 3 |
| GO:0090575 | RNA polymerase II transcription regulator complex | 10/187 | 274/22283 | 0.000112303 | 0.003357874 | 0.002919891 | JUND/RUNX3/TAF10/STAT3/NFIL3/CHD4/MAFF/CEBPZ/MED23/ASCC3 | 10 |
| GO:0000307 | cyclin-dependent protein kinase holoenzyme complex | 4/187 | 51/22283 | 0.000882105 | 0.023977211 | 0.020849748 | CCNL1/CDK13/CCNK/CDK12 | 4 |
| GO:0000123 | histone acetyltransferase complex | 5/187 | 98/22283 | 0.001426525 | 0.035544249 | 0.030908043 | TAF10/EPC1/BRD1/KAT6B/ING3 | 5 |
| GO:0032806 | carboxy-terminal domain protein kinase complex | 3/187 | 28/22283 | 0.001632531 | 0.037548217 | 0.032650623 | CDK13/CCNK/CDK12 | 3 |
| GO:0031248 | protein acetyltransferase complex | 5/187 | 108/22283 | 0.002189419 | 0.043122607 | 0.03749792 | TAF10/EPC1/BRD1/KAT6B/ING3 | 5 |
| GO:1902493 | acetyltransferase complex | 5/187 | 108/22283 | 0.002189419 | 0.043122607 | 0.03749792 | TAF10/EPC1/BRD1/KAT6B/ING3 | 5 |
| GO:0042571 | immunoglobulin complex, circulating | 4/187 | 66/22283 | 0.002307564 | 0.043122607 | 0.03749792 | IGHA1/IGLC3/IGKC/IGLC2 | 4 |

### Supplementary Table 6. GO pathways Monocytes.

#### Biological pathways

| ID | Description | GeneRatio | BgRatio | pvalue | p.adjust | qvalue | geneID | Count |
|---|---|---|---|---|---|---|---|---|
| GO:0031638 | zymogen activation | 7/177 | 72/21069 | 2.44E-06 | 0.006707158 | 0.005773932 | HP/ASPH/THBS1/THBD/VSIR/RUNX1/CTSL | 7 |
| GO:0034976 | response to endoplasmic reticulum stress | 12/177 | 295/21069 | 7.85E-06 | 0.010799681 | 0.009297027 | CLU/CXCL8/UFD1/RACK1/PTPN1/SELENOK/THBS1/NIBAN1/PPP1R15A/SELENOS/PIK3R1/UBE2J1 | 12 |
| GO:1990748 | cellular detoxification | 8/177 | 130/21069 | 1.44E-05 | 0.013191375 | 0.011355944 | HBB/HP/SELENOW/PIM1/HBA2/SELENOS/ALOX5AP/SELENOT | 8 |
| GO:0097237 | cellular response to toxic substance | 8/177 | 145/21069 | 3.17E-05 | 0.021191257 | 0.018242731 | HBB/HP/SELENOW/PIM1/HBA2/SELENOS/ALOX5AP/SELENOT | 8 |
| GO:0061097 | regulation of protein tyrosine kinase activity | 7/177 | 111/21069 | 4.27E-05 | 0.021191257 | 0.018242731 | HBEGF/AREG/EREG/RACK1/PTPN1/SH3BP5/FCGR1A | 7 |
| GO:0098869 | cellular oxidant detoxification | 7/177 | 115/21069 | 5.36E-05 | 0.021191257 | 0.018242731 | HBB/HP/SELENOW/HBA2/SELENOS/ALOX5AP/SELENOT | 7 |
| GO:0006911 | phagocytosis, engulfment | 7/177 | 123/21069 | 8.21E-05 | 0.021191257 | 0.018242731 | IGHA1/IGLC3/IGLC2/IGKC/THBS1/FCGR1A/RAB31 | 7 |
| GO:1901654 | response to ketone | 9/177 | 215/21069 | 8.84E-05 | 0.021191257 | 0.018242731 | F5/THBS1/KLF9/PTGER2/AHR/FOXO3/ATP2B1/SGK1/PTAFR | 9 |
| GO:1903573 | negative regulation of response to endoplasmic reticulum stress | 5/177 | 54/21069 | 8.96E-05 | 0.021191257 | 0.018242731 | CLU/RACK1/PTPN1/PPP1R15A/SELENOS | 5 |
| GO:0098532 | histone H3-K27 trimethylation | 3/177 | 11/21069 | 9.15E-05 | 0.021191257 | 0.018242731 | H1-4/H1-2/H1-3 | 3 |
| GO:0098754 | detoxification | 8/177 | 170/21069 | 9.76E-05 | 0.021191257 | 0.018242731 | HBB/HP/SELENOW/PIM1/HBA2/SELENOS/ALOX5AP/SELENOT | 8 |
| GO:0042149 | cellular response to glucose starvation | 5/177 | 55/21069 | 9.79E-05 | 0.021191257 | 0.018242731 | UPP1/PLIN2/FOXO3/PLIN3/CPEB4 | 5 |
| GO:0016485 | protein processing | 10/177 | 275/21069 | 0.000115714 | 0.021191257 | 0.018242731 | HP/ASPH/MAFB/THBS1/THBD/VSIR/CPD/RUNX1/CST7/CTSL | 10 |
| GO:0097193 | intrinsic apoptotic signaling pathway | 11/177 | 332/21069 | 0.000120481 | 0.021191257 | 0.018242731 | CLU/BCL2A1/HIF1A/RACK1/PTPN1/SELENOK/PPP1R15A/SELENOS/PIK3R1/CDKN2D/BCLAF1 | 11 |
| GO:0006959 | humoral immune response | 12/177 | 392/21069 | 0.000124824 | 0.021191257 | 0.018242731 | CLU/IGHA1/RGCC/CXCL8/CXCL2/IGLC3/IGLC2/CR1/IGKC/CXCL3/GPR183/TREM1 | 12 |
| GO:0099024 | plasma membrane invagination | 7/177 | 132/21069 | 0.000127938 | 0.021191257 | 0.018242731 | IGHA1/IGLC3/IGLC2/IGKC/THBS1/FCGR1A/RAB31 | 7 |
| GO:0045860 | positive regulation of protein kinase activity | 13/177 | 457/21069 | 0.000135456 | 0.021191257 | 0.018242731 | CLU/RGCC/HBEGF/AREG/EREG/FAM20A/PTPN1/THBS1/ACSL1/DIPK2A/SYAP1/ADAM9/FCGR1A | 13 |
| GO:0042542 | response to hydrogen peroxide | 7/177 | 134/21069 | 0.000140504 | 0.021191257 | 0.018242731 | HBB/HP/AREG/RACK1/HBA2/FOXO3/ADAM9 | 7 |
| GO:0045741 | positive regulation of epidermal growth factor-activated receptor activity | 3/177 | 13/21069 | 0.000156704 | 0.021191257 | 0.018242731 | HBEGF/AREG/EREG | 3 |
| GO:0071389 | cellular response to mineralocorticoid stimulus | 3/177 | 13/21069 | 0.000156704 | 0.021191257 | 0.018242731 | FOXO3/ATP2B1/SGK1 | 3 |
| GO:1905897 | regulation of response to endoplasmic reticulum stress | 6/177 | 98/21069 | 0.00017938 | 0.021191257 | 0.018242731 | CLU/RACK1/PTPN1/PPP1R15A/SELENOS/PIK3R1 | 6 |
| GO:1900101 | regulation of endoplasmic reticulum unfolded protein response | 4/177 | 34/21069 | 0.000183371 | 0.021191257 | 0.018242731 | RACK1/PTPN1/PPP1R15A/PIK3R1 | 4 |
| GO:0010324 | membrane invagination | 7/177 | 141/21069 | 0.000192602 | 0.021191257 | 0.018242731 | IGHA1/IGLC3/IGLC2/IGKC/THBS1/FCGR1A/RAB31 | 7 |
| GO:0032640 | tumor necrosis factor production | 9/177 | 239/21069 | 0.000196296 | 0.021191257 | 0.018242731 | CLU/SELENOK/THBS1/VSIR/SELENOS/PIK3R1/MAPKAPK2/UBE2J1/PTAFR | 9 |
| GO:0032680 | regulation of tumor necrosis factor production | 9/177 | 239/21069 | 0.000196296 | 0.021191257 | 0.018242731 | CLU/SELENOK/THBS1/VSIR/SELENOS/PIK3R1/MAPKAPK2/UBE2J1/PTAFR | 9 |
| GO:0019915 | lipid storage | 6/177 | 100/21069 | 0.000200354 | 0.021191257 | 0.018242731 | NRIP1/SQLE/GM2A/PLIN2/PLIN3/EHD1 | 6 |
| GO:0006956 | complement activation | 7/177 | 145/21069 | 0.000228754 | 0.0216314 | 0.018621634 | CLU/IGHA1/RGCC/IGLC3/IGLC2/CR1/IGKC | 7 |
| GO:0071706 | tumor necrosis factor superfamily cytokine production | 9/177 | 244/21069 | 0.000228938 | 0.0216314 | 0.018621634 | CLU/SELENOK/THBS1/VSIR/SELENOS/PIK3R1/MAPKAPK2/UBE2J1/PTAFR | 9 |
| GO:1903555 | regulation of tumor necrosis factor superfamily cytokine production | 9/177 | 244/21069 | 0.000228938 | 0.0216314 | 0.018621634 | CLU/SELENOK/THBS1/VSIR/SELENOS/PIK3R1/MAPKAPK2/UBE2J1/PTAFR | 9 |
| GO:0031669 | cellular response to nutrient levels | 9/177 | 245/21069 | 0.000235979 | 0.0216314 | 0.018621634 | PIM1/UPP1/PLIN2/FOXO3/ATP2B1/PLIN3/CPEB4/GABARAPL1/MAP1LC3B | 9 |
| GO:0061098 | positive regulation of protein tyrosine kinase activity | 5/177 | 67/21069 | 0.000250631 | 0.022233401 | 0.019139874 | HBEGF/AREG/EREG/PTPN1/FCGR1A | 5 |
| GO:0035966 | response to topologically incorrect protein | 8/177 | 198/21069 | 0.000277672 | 0.023862422 | 0.020542234 | CLU/UFD1/RACK1/PTPN1/THBS1/PPP1R15A/SELENOS/PIK3R1 | 8 |
| GO:0097306 | cellular response to alcohol | 6/177 | 107/21069 | 0.00028942 | 0.023946677 | 0.020614766 | KLF9/PTGER2/AHR/FOXO3/ATP2B1/SGK1 | 6 |
| GO:0050864 | regulation of B cell activation | 8/177 | 200/21069 | 0.000297121 | 0.023946677 | 0.020614766 | IGHA1/IGLC3/IGLC2/CR1/IGKC/GPR183/SAMSN1/AHR | 8 |
| GO:0002237 | response to molecule of bacterial origin | 12/177 | 432/21069 | 0.000304776 | 0.023946677 | 0.020614766 | CXCL8/CXCL2/TRIB1/CXCL3/THBD/PTGER2/SELENOS/MAPKAPK2/C5AR1/NLRP3/ADAM9/PTAFR | 12 |
| GO:0006958 | complement activation, classical pathway | 6/177 | 111/21069 | 0.000352735 | 0.026896826 | 0.023154435 | CLU/IGHA1/IGLC3/IGLC2/CR1/IGKC | 6 |
| GO:1901655 | cellular response to ketone | 6/177 | 112/21069 | 0.00037014 | 0.026896826 | 0.023154435 | KLF9/PTGER2/AHR/FOXO3/ATP2B1/SGK1 | 6 |
| GO:0048511 | rhythmic process | 10/177 | 318/21069 | 0.000371665 | 0.026896826 | 0.023154435 | NFIL3/PROK2/EREG/NRIP1/RACK1/KLF9/NAMPT/AHR/FOXO3/BHLHE40 | 10 |
| GO:0009749 | response to glucose | 8/177 | 210/21069 | 0.000411809 | 0.029037815 | 0.024997529 | HIF1A/RACK1/THBS1/SELENOS/PIM3/CCDC186/SELENOT/FOXO3 | 8 |
| GO:0033002 | muscle cell proliferation | 9/177 | 266/21069 | 0.000430966 | 0.029075023 | 0.02502956 | HBEGF/TRIB1/EREG/PIM1/THBS1/RUNX1/DIPK2A/JARID2/PTAFR | 9 |
| GO:0038128 | ERBB2 signaling pathway | 3/177 | 18/21069 | 0.000433482 | 0.029075023 | 0.02502956 | HBEGF/AREG/EREG | 3 |
| GO:0000302 | response to reactive oxygen species | 8/177 | 214/21069 | 0.000466744 | 0.030560622 | 0.026308455 | HBB/HP/AREG/HIF1A/RACK1/HBA2/FOXO3/ADAM9 | 8 |
| GO:1900102 | negative regulation of endoplasmic reticulum unfolded protein response | 3/177 | 19/21069 | 0.00051159 | 0.032717933 | 0.028165601 | RACK1/PTPN1/PPP1R15A | 3 |
| GO:0050900 | leukocyte migration | 12/177 | 459/21069 | 0.000523839 | 0.032739937 | 0.028184543 | CXCL8/CXCL2/CXCL3/GPR183/SELENOK/THBS1/PIK3R1/C5AR1/CCR1/SBDS/TREM1/PTAFR | 12 |
| GO:0032496 | response to lipopolysaccharide | 11/177 | 398/21069 | 0.000565988 | 0.033603991 | 0.028928373 | CXCL8/CXCL2/TRIB1/CXCL3/THBD/PTGER2/SELENOS/MAPKAPK2/NLRP3/ADAM9/PTAFR | 11 |
| GO:0030217 | T cell differentiation | 10/177 | 336/21069 | 0.000570971 | 0.033603991 | 0.028928373 | CR1/HLX/MAFB/GPR183/VSIR/PIK3R1/RUNX1/FOXO3/NLRP3/CTSL | 10 |
| GO:0006909 | phagocytosis | 10/177 | 337/21069 | 0.000584232 | 0.033603991 | 0.028928373 | IGHA1/IGLC3/IGLC2/IGKC/RACK1/THBS1/LDLR/DYSF/FCGR1A/RAB31 | 10 |
| GO:0009746 | response to hexose | 8/177 | 222/21069 | 0.000594479 | 0.033603991 | 0.028928373 | HIF1A/RACK1/THBS1/SELENOS/PIM3/CCDC186/SELENOT/FOXO3 | 8 |
| GO:0001678 | cellular glucose homeostasis | 7/177 | 171/21069 | 0.000618845 | 0.033603991 | 0.028928373 | HIF1A/RACK1/PIK3R1/PIM3/CCDC186/SELENOT/FOXO3 | 7 |

| ID | Description | GeneRatio | BgRatio | pvalue | p.adjust | qvalue | geneID | Count |
|---|---|---|---|---|---|---|---|---|
| GO:0030595 | leukocyte chemotaxis | 9/177 | 280/21069 | 0.000623201 | 0.033603991 | 0.028928373 | CXCL8/CXCL2/CXCL3/GPR183/THBS1/C5AR1/CCR1/SBDS/TREM1 | 9 |
| GO:0097529 | myeloid leukocyte migration | 9/177 | 280/21069 | 0.000623201 | 0.033603991 | 0.028928373 | CXCL8/CXCL2/CXCL3/SELENOK/THBS1/PIK3R1/C5AR1/CCR1/TREM1 | 9 |
| GO:0030968 | endoplasmic reticulum unfolded protein response | 5/177 | 82/21069 | 0.000639917 | 0.033841777 | 0.029133074 | RACK1/PTPN1/PPP1R15A/SELENOS/PIK3R1 | 5 |
| GO:0031668 | cellular response to extracellular stimulus | 9/177 | 283/21069 | 0.000672378 | 0.03390963 | 0.029191486 | PIM1/UPP1/PLIN2/FOXO3/ATP2B1/PLIN3/CPEB4/GABARAPL1/MAP1LC3B | 9 |
| GO:0097530 | granulocyte migration | 7/177 | 174/21069 | 0.000685951 | 0.03390963 | 0.029191486 | CXCL8/CXCL2/CXCL3/SELENOK/THBS1/C5AR1/TREM1 | 7 |
| GO:0002274 | myeloid leukocyte activation | 9/177 | 285/21069 | 0.000706892 | 0.03390963 | 0.029191486 | CLU/CXCL8/THBS1/LDLR/C5AR1/DYSF/CST7/ADAM9/PTAFR | 9 |
| GO:1902107 | positive regulation of leukocyte differentiation | 8/177 | 228/21069 | 0.000707722 | 0.03390963 | 0.029191486 | TRIB1/CR1/HLX/VSIR/RUNX1/FOXO3/CCR1/NLRP3 | 8 |
| GO:1903708 | positive regulation of hemopoiesis | 8/177 | 228/21069 | 0.000707722 | 0.03390963 | 0.029191486 | TRIB1/CR1/HLX/VSIR/RUNX1/FOXO3/CCR1/NLRP3 | 8 |
| GO:0009636 | response to toxic substance | 9/177 | 286/21069 | 0.000724683 | 0.03390963 | 0.029191486 | HBB/HP/SELENOW/PIM1/HBA2/SELENOS/AHR/ALOX5AP/SELENOT | 9 |
| GO:0034284 | response to monosaccharide | 8/177 | 230/21069 | 0.000749114 | 0.03390963 | 0.029191486 | HIF1A/RACK1/THBS1/SELENOS/PIM3/CCDC186/SELENOT/FOXO3 | 8 |
| GO:2000273 | positive regulation of signaling receptor activity | 4/177 | 49/21069 | 0.000759568 | 0.03390963 | 0.029191486 | HBEGF/AREG/EREG/HIF1A | 4 |
| GO:1902106 | negative regulation of leukocyte differentiation | 6/177 | 129/21069 | 0.000782782 | 0.03390963 | 0.029191486 | TRIB1/CR1/HLX/MAFB/PIK3R1/RUNX1 | 6 |
| GO:0007171 | activation of transmembrane receptor protein tyrosine kinase activity | 3/177 | 22/21069 | 0.000798133 | 0.03390963 | 0.029191486 | HBEGF/AREG/EREG | 3 |
| GO:0030728 | ovulation | 3/177 | 22/21069 | 0.000798133 | 0.03390963 | 0.029191486 | EREG/NRIP1/FOXO3 | 3 |
| GO:0071496 | cellular response to external stimulus | 10/177 | 351/21069 | 0.000798325 | 0.03390963 | 0.029191486 | PIM1/UPP1/GADD45A/PLIN2/FOXO3/ATP2B1/PLIN3/CPEB4/GABARAPL1/MAP1LC3B | 10 |
| GO:0002687 | positive regulation of leukocyte migration | 7/177 | 179/21069 | 0.0008105 | 0.03390963 | 0.029191486 | CXCL8/SELENOK/THBS1/PIK3R1/C5AR1/CCR1/PTAFR | 7 |
| GO:2001233 | regulation of apoptotic signaling pathway | 11/177 | 416/21069 | 0.000813831 | 0.03390963 | 0.029191486 | CLU/G0S2/HIF1A/RACK1/PTPN1/THBS1/LMNA/SELENOS/CDKN2D/BCLAF1/ITPRIP | 11 |
| GO:0002526 | acute inflammatory response | 6/177 | 131/21069 | 0.000848296 | 0.03447684 | 0.029679776 | HP/SELENOS/ALOX5AP/TREM1/NLRP3/FCGR1A | 6 |
| GO:0050730 | regulation of peptidyl-tyrosine phosphorylation | 9/177 | 293/21069 | 0.000859735 | 0.03447684 | 0.029679776 | HBEGF/AREG/EREG/RACK1/EHD4/PTPN1/SAMSN1/SH3BP5/FCGR1A | 9 |
| GO:0031960 | response to corticosteroid | 7/177 | 181/21069 | 0.000865055 | 0.03447684 | 0.029679776 | AREG/KLF9/FOXO3/ATP2B1/SGK1/ADAM9/PTAFR | 7 |
| GO:0080182 | histone H3-K4 trimethylation | 3/177 | 23/21069 | 0.000912208 | 0.035554836 | 0.03060778 | H1-4/H1-2/H1-3 | 3 |
| GO:0032760 | positive regulation of tumor necrosis factor production | 6/177 | 133/21069 | 0.000917961 | 0.035554836 | 0.03060778 | CLU/SELENOK/THBS1/PIK3R1/MAPKAPK2/PTAFR | 6 |
| GO:2001242 | regulation of intrinsic apoptotic signaling pathway | 7/177 | 185/21069 | 0.000982847 | 0.037368023 | 0.032168683 | CLU/HIF1A/RACK1/PTPN1/SELENOS/CDKN2D/BCLAF1 | 7 |
| GO:1903707 | negative regulation of hemopoiesis | 6/177 | 135/21069 | 0.000991951 | 0.037368023 | 0.032168683 | TRIB1/CR1/HLX/MAFB/PIK3R1/RUNX1 | 6 |
| GO:0051604 | protein maturation | 10/177 | 362/21069 | 0.001008631 | 0.037482924 | 0.032267597 | HP/ASPH/MAFB/THBS1/THBD/VSIR/CPD/RUNX1/CST7/CTSL | 10 |
| GO:0019755 | one-carbon compound transport | 3/177 | 24/21069 | 0.001036115 | 0.037990888 | 0.032704883 | HBB/AQP9/HBA2 | 3 |
| GO:1903557 | positive regulation of tumor necrosis factor superfamily cytokine production | 6/177 | 137/21069 | 0.001070444 | 0.038733161 | 0.033343877 | CLU/SELENOK/THBS1/PIK3R1/MAPKAPK2/PTAFR | 6 |
| GO:0002455 | humoral immune response mediated by circulating immunoglobulin | 6/177 | 138/21069 | 0.001111435 | 0.039156463 | 0.033708281 | CLU/IGHA1/IGLC3/IGLC2/CR1/IGKC | 6 |
| GO:0009267 | cellular response to starvation | 7/177 | 189/21069 | 0.00111292 | 0.039156463 | 0.033708281 | UPP1/PLIN2/FOXO3/PLIN3/CPEB4/GABARAPL1/MAP1LC3B | 7 |
| GO:0071900 | regulation of protein serine/threonine kinase activity | 11/177 | 433/21069 | 0.001124858 | 0.039156463 | 0.033708281 | RGCC/TRIB1/FAM20A/PTPN1/THBS1/GADD45A/ACSL1/DIPK2A/SYAP1/CDKN2D/ADAM9 | 11 |
| GO:0042116 | macrophage activation | 6/177 | 139/21069 | 0.001153619 | 0.039541802 | 0.034040005 | CLU/THBS1/LDLR/C5AR1/DYSF/CST7 | 6 |
| GO:0060326 | cell chemotaxis | 10/177 | 369/21069 | 0.001164686 | 0.039541802 | 0.034040005 | CXCL8/CXCL2/HBEGF/CXCL3/GPR183/THBS1/C5AR1/CCR1/SBDS/TREM1 | 10 |
| GO:0051591 | response to cAMP | 5/177 | 94/21069 | 0.001187427 | 0.03982225 | 0.034281431 | AREG/AQP9/THBD/AHR/PTAFR | 5 |
| GO:0071902 | positive regulation of protein serine/threonine kinase activity | 8/177 | 249/21069 | 0.001248046 | 0.041350911 | 0.035597397 | RGCC/FAM20A/PTPN1/THBS1/ACSL1/DIPK2A/SYAP1/ADAM9 | 8 |
| GO:0030502 | negative regulation of bone mineralization | 3/177 | 26/21069 | 0.001314674 | 0.043039926 | 0.037051404 | RFLNB/HIF1A/CCR1 | 3 |
| GO:0006910 | phagocytosis, recognition | 5/177 | 97/21069 | 0.001366287 | 0.04420339 | 0.038052985 | IGHA1/IGLC3/IGLC2/IGKC/FCGR1A | 5 |
| GO:0071621 | granulocyte chemotaxis | 6/177 | 145/21069 | 0.001433091 | 0.045340733 | 0.039032079 | CXCL8/CXCL2/CXCL3/THBS1/C5AR1/TREM1 | 6 |
| GO:0018023 | peptidyl-lysine trimethylation | 4/177 | 58/21069 | 0.001434416 | 0.045340733 | 0.039032079 | H1-4/H1-2/H1-3/LMNA | 4 |
| GO:0070734 | histone H3-K27 methylation | 3/177 | 27/21069 | 0.001469927 | 0.045353103 | 0.039042728 | H1-4/H1-2/H1-3 | 3 |
| GO:0045582 | positive regulation of T cell differentiation | 6/177 | 146/21069 | 0.001484283 | 0.045353103 | 0.039042728 | CR1/HLX/VSIR/RUNX1/FOXO3/NLRP3 | 6 |
| GO:1990266 | neutrophil migration | 6/177 | 146/21069 | 0.001484283 | 0.045353103 | 0.039042728 | CXCL8/CXCL2/CXCL3/SELENOK/C5AR1/TREM1 | 6 |
| GO:1902105 | regulation of leukocyte differentiation | 10/177 | 382/21069 | 0.001506785 | 0.045534721 | 0.039199077 | TRIB1/CR1/HLX/MAFB/VSIR/PIK3R1/RUNX1/FOXO3/CCR1/NLRP3 | 10 |
| GO:0009743 | response to carbohydrate | 8/177 | 257/21069 | 0.001524647 | 0.045573688 | 0.039232622 | HIF1A/RACK1/THBS1/SELENOS/PIM3/CCDC186/SELENOT/FOXO3 | 8 |
| GO:0043367 | CD4-positive, alpha-beta T cell differentiation | 5/177 | 100/21069 | 0.001564277 | 0.0462555 | 0.039819567 | HLX/GPR183/RUNX1/NLRP3/CTSL | 5 |
| GO:0072593 | reactive oxygen species metabolic process | 8/177 | 259/21069 | 0.001600874 | 0.046834069 | 0.040317635 | HBB/HP/HIF1A/HBA2/THBS1/GADD45A/FOXO3/STK17A | 8 |
| GO:0035710 | CD4-positive, alpha-beta T cell activation | 6/177 | 149/21069 | 0.001646251 | 0.047158236 | 0.040596698 | HLX/GPR183/VSIR/RUNX1/NLRP3/CTSL | 6 |
| GO:0071333 | cellular response to glucose stimulus | 6/177 | 149/21069 | 0.001646251 | 0.047158236 | 0.040596698 | HIF1A/RACK1/PIM3/CCDC186/SELENOT/FOXO3 | 6 |
| GO:2001234 | negative regulation of apoptotic signaling pathway | 8/177 | 262/21069 | 0.001720847 | 0.048427006 | 0.041688933 | CLU/HIF1A/PTPN1/THBS1/LMNA/SELENOS/CDKN2D/ITPRIP | 8 |
| GO:0002673 | regulation of acute inflammatory response | 4/177 | 61/21069 | 0.001730262 | 0.048427006 | 0.041688933 | SELENOS/ALOX5AP/NLRP3/FCGR1A | 4 |
| GO:0071331 | cellular response to hexose stimulus | 6/177 | 151/21069 | 0.001761466 | 0.048427006 | 0.041688933 | HIF1A/RACK1/PIM3/CCDC186/SELENOT/FOXO3 | 6 |
| GO:0010714 | positive regulation of collagen metabolic process | 3/177 | 29/21069 | 0.001813811 | 0.048427006 | 0.041688933 | RGCC/VSIR/RUNX1 | 3 |
| GO:0010884 | positive regulation of lipid storage | 3/177 | 29/21069 | 0.001813811 | 0.048427006 | 0.041688933 | PLIN2/PLIN3/EHD1 | 3 |
| GO:0030970 | retrograde protein transport, ER to cytosol | 3/177 | 29/21069 | 0.001813811 | 0.048427006 | 0.041688933 | UFD1/SELENOS/UBE2J1 | 3 |
| GO:1903513 | endoplasmic reticulum to cytosol transport | 3/177 | 29/21069 | 0.001813811 | 0.048427006 | 0.041688933 | UFD1/SELENOS/UBE2J1 | 3 |
| GO:0070098 | chemokine-mediated signaling pathway | 5/177 | 104/21069 | 0.001860098 | 0.049185288 | 0.042341708 | CXCL8/CXCL2/CXCL3/HIF1A/CCR1 | 5 |
| GO:0071326 | cellular response to monosaccharide stimulus | 6/177 | 153/21069 | 0.0018827 | 0.049308811 | 0.042448045 | HIF1A/RACK1/PIM3/CCDC186/SELENOT/FOXO3 | 6 |

### Molecular function

| ID | Description | GeneRatio | BgRatio | pvalue | p.adjust | qvalue | geneID | Count |
|---|---|---|---|---|---|---|---|---|
| GO:0016209 | antioxidant activity | 7/180 | 99/20758 | 2.50E-05 | 0.005193765 | 0.004731162 | HBB/HP/SELENOW/HBA2/SELENOS/ALOX5AP/SELENOT | 7 |
| GO:0019207 | kinase regulator activity | 11/180 | 270/20758 | 2.50E-05 | 0.005193765 | 0.004731162 | RGCC/HBEGF/TRIB1/AREG/EREG/FAM20A/RACK1/SH3BP5/PIK3R1/CDKN2D/ITPRIP | 11 |
| GO:0019887 | protein kinase regulator activity | 10/180 | 239/20758 | 4.69E-05 | 0.006503054 | 0.005923835 | RGCC/HBEGF/TRIB1/AREG/EREG/FAM20A/RACK1/SH3BP5/CDKN2D/ITPRIP | 10 |
| GO:0030297 | transmembrane receptor protein tyrosine kinase activator activity | 3/180 | 16/20758 | 0.000330445 | 0.034366232 | 0.031305272 | HBEGF/AREG/EREG | 3 |
| GO:0045236 | CXCR chemokine receptor binding | 3/180 | 19/20758 | 0.000560945 | 0.040696752 | 0.03707194 | CXCL8/CXCL2/CXCL3 | 3 |
| GO:0004860 | protein kinase inhibitor activity | 5/180 | 78/20758 | 0.000586972 | 0.040696752 | 0.03707194 | TRIB1/RACK1/SH3BP5/CDKN2D/ITPRIP | 5 |
| GO:0019210 | kinase inhibitor activity | 5/180 | 83/20758 | 0.000779663 | 0.046334231 | 0.042207296 | TRIB1/RACK1/SH3BP5/CDKN2D/ITPRIP | 5 |

### Cellular components

| ID | Description | GeneRatio | BgRatio | pvalue | p.adjust | qvalue | geneID | Count |
|---|---|---|---|---|---|---|---|---|
| GO:0071682 | endocytic vesicle lumen | 4/181 | 25/22283 | 4.66E-05 | 0.005600553 | 0.004873748 | HBB/HP/HBA2/CTSL | 4 |
| GO:0071735 | IgG immunoglobulin complex | 3/181 | 10/22283 | 6.07E-05 | 0.005600553 | 0.004873748 | IGHA1/IGLC3/IGKC | 3 |
| GO:0071745 | IgA immunoglobulin complex | 3/181 | 10/22283 | 6.07E-05 | 0.005600553 | 0.004873748 | IGHA1/IGLC3/IGKC | 3 |
| GO:0031838 | haptoglobin-hemoglobin complex | 3/181 | 11/22283 | 8.29E-05 | 0.005741053 | 0.004996014 | HBB/HP/HBA2 | 3 |
| GO:0072562 | blood microparticle | 8/181 | 181/22283 | 0.000120154 | 0.006091421 | 0.005300914 | CLU/HBB/IGHA1/HP/IGLC3/IGLC2/IGKC/HBA2 | 8 |
| GO:0030139 | endocytic vesicle | 13/181 | 471/22283 | 0.000131944 | 0.006091421 | 0.005300914 | HBB/HP/HBEGF/AREG/EREG/EHD4/HBA2/LDLR/DYSF/EHD1/CTSL/FCGR1A/RAB31 | 13 |
| GO:0070820 | tertiary granule | 8/181 | 216/22283 | 0.000399004 | 0.01578916 | 0.013740139 | HBB/HP/CR1/SLC2A3/MCEMP1/CYSTM1/FCAR/PTAFR | 8 |
| GO:0008023 | transcription elongation factor complex | 4/181 | 51/22283 | 0.000781219 | 0.027049711 | 0.023539365 | EAF1/ELOB/ELOC/AFF4 | 4 |
| GO:0000786 | nucleosome | 6/181 | 137/22283 | 0.000902323 | 0.027771486 | 0.024167472 | H1-4/H1-2/H2AC20/H1-3/H2AJ/EPC1 | 6 |
| GO:0030667 | secretory granule membrane | 10/181 | 380/22283 | 0.001133307 | 0.031392609 | 0.027318668 | CR1/SLC2A3/CKAP4/MCEMP1/ANPEP/C5AR1/CYSTM1/FCAR/PTAFR/RAB31 | 10 |
| GO:0000791 | euchromatin | 4/181 | 61/22283 | 0.001531823 | 0.037583269 | 0.032705942 | H1-2/H1-3/HIF1A/AFF4 | 4 |
| GO:0005793 | endoplasmic reticulum-Golgi intermediate compartment | 6/181 | 157/22283 | 0.001815248 | 0.037583269 | 0.032705942 | AREG/F5/ANPEP/ZDHHC20/MGAT4A/TMED5 | 6 |
| GO:0005811 | lipid droplet | 5/181 | 107/22283 | 0.001823376 | 0.037583269 | 0.032705942 | G0S2/CKAP4/PLIN2/PLIN3/EHD1 | 5 |
| GO:0070821 | tertiary granule membrane | 5/181 | 108/22283 | 0.001899515 | 0.037583269 | 0.032705942 | SLC2A3/MCEMP1/CYSTM1/FCAR/PTAFR | 5 |
| GO:0042571 | immunoglobulin complex, circulating | 4/181 | 66/22283 | 0.002050113 | 0.037858755 | 0.032945677 | IGHA1/IGLC3/IGLC2/IGKC | 4 |

#### Supplementary Table 7. KEGG pathways

| ID | Description | GeneRatio | BgRatio | pvalue | p.adjust | qvalue | geneID | Count |
|---|---|---|---|---|---|---|---|---|
| hsa03082 | ATP-dependent chromatin remodeling | Sep-83 | 117/8579 | 1.69E-06 | 0.00034992 | 0.000274029 | 8338/29117/80314/1108/1107/6595/54556/6602/54617 | 9 |
| hsa05211 | Renal cell carcinoma | Jul-83 | 69/8579 | 4.08E-06 | 0.00042273 | 0.000331047 | 6923/5295/5062/6921/2889/673/6655 | 7 |
| hsa04062 | Chemokine signaling pathway | Sep-83 | 192/8579 | 9.20E-05 | 0.006345323 | 0.004969132 | 7852/5295/4792/6774/1794/57580/5336/673/6655 | 9 |
| hsa04012 | ErbB signaling pathway | Jun-83 | 85/8579 | 0.000162446 | 0.008406566 | 0.006583327 | 374/5295/5062/5336/673/6655 | 6 |
| hsa05223 | Non-small cell lung cancer | May-83 | 72/8579 | 0.000632105 | 0.026169149 | 0.020493511 | 5295/6774/5336/673/6655 | 5 |
| hsa01521 | EGFR tyrosine kinase inhibitor resistance | May-83 | 79/8579 | 0.000965856 | 0.028572121 | 0.02237532 | 5295/6774/5336/673/6655 | 5 |
| hsa04722 | Neurotrophin signaling pathway | Jun-83 | 119/8579 | 0.000992935 | 0.028572121 | 0.02237532 | 5295/4792/2889/5336/673/6655 | 6 |
| hsa05225 | Hepatocellular carcinoma | Jul-83 | 168/8579 | 0.001159585 | 0.028572121 | 0.02237532 | 5295/29117/6595/5336/673/6602/6655 | 7 |
| hsa04141 | Protein processing in endoplasmic reticulum | Jul-83 | 170/8579 | 0.001242266 | 0.028572121 | 0.02237532 | 3309/55829/7353/6238/23645/4121/8720 | 7 |
| hsa05162 | Measles | Jun-83 | 138/8579 | 0.002125798 | 0.044004014 | 0.034460301 | 7128/10399/5295/4792/6774/5610 | 6 |
